# Usage of an Alternative Translation Start Site in *PRKN* mutation carriers: Key to Later-Onset Parkinson’s Disease and a Novel Therapeutic Target

**DOI:** 10.1101/2025.05.07.25326886

**Authors:** Arian Hach, Katja Lohmann, Manabu Funayama, Poornima Jayadev Menon, Eva-Juliane Vollstedt, Teresa Kleinz, Hiroyo Yoshino, Suzanne Lesage, Britta Meier, Carolyn Mary Sue, Jean-Christophe Corvol, Alexis Brice, Nobutaka Hattori, Christine Klein, Aleksandar Rakovic

## Abstract

Biallelic pathogenic variants of *PRKN,* encoding the Parkin RBR E3 ubiquitin protein ligase, are the most common known cause of autosomal recessive Parkinson’s disease (PD). PARK-*PRKN* is characterized by an early median age at onset (AAO) of 31 years with a wide range (3-81 years). When evaluating the 16 previously published carriers of a homozygous deletion of Exon 2 (*PRKN*^delEx2^) from the MDSGene database, the median AAO is later (39.5 years; range: 25-75 years) than in carriers of other *PRKN* pathogenic variants. Here, we investigated 26 homozygous *PRKN*^delEx2^ patients, including 20 from additional sources, and confirmed the later median AAO (37 years, range: 13-60 years). Furthermore, we investigated one carrier who was still unaffected in her 60s. To elucidate the functional basis for this observation, we used induced pluripotent stem cell (iPSC)-derived dopaminergic neurons (iDN) from this unaffected carrier as well as genome-edited iDNs from two control lines and neuroblastoma cell lines (SH-SY5Y) with the introduction of a homozygous *PRKN*^delEx2^. We observed elevated levels of an N-terminally truncated form of Parkin (Parkin^Δ1–79)^, starting at an internal translation initiation site (TIS) in Exon 3 (p.Met80 in the full-length protein) in cells with the Exon 2 deletion. Furthermore, *in silico* prediction and Parkin quantification in isogenic neuroblastoma cell lines suggested the presence of residual Parkin^Δ1–79^ also for other *PRKN* variants upstream of the alternative TIS. iDNs from *PRKN*^delEx2^ carriers partially retained Parkin E3 ubiquitin ligase activity in contrast to carriers of other exonic deletions in *PRKN* (*PRKN*^delEx3^, *PRKN*^delEx7^) via the expression of Parkin^Δ1–79^. Importantly, endogenous Parkin E3 ubiquitin ligase activity of Parkin^Δ1–79^ was enhanced in neurons derived from a homozygous *PRKN*^delEx2^ variant carrier upon treatment with a small molecule allosteric modulator of Parkin (BIO-2007817). In summary, the later AAO in patients with homozygous *PRKN*^delEx2^ is associated with partially retained Parkin function. The residual ligase activity can be further increased pharmacologically, providing mutation-specific personalized counseling opportunities and a potential novel therapy for selected patients with PARK-*PRKN*.

## Introduction

While the majority of PD patients are classified as idiopathic with an unknown origin, monogenic causes for PD constitute up to 15% of all patients.^1, 2^ Of these, pathogenic variants in the *PRKN* gene are the most common cause of familial, monogenic autosomal recessive PD (PARK-*PRKN*).^1, 2^

PD is typically characterized by a relatively late median age at disease onset (AAO) of 60 years, with increasing prevalence at higher ages.^3^ Patients with a monogenic cause of PD usually develop the disease at an earlier AAO. For instance, the median AAO for PARK-*PRKN* is 31 years, notably, with a range from 3-81 years.^4–6^ Although no variant-specific genotype-phenotype correlation within PARK-*PRKN* patients has been detected yet, there is a 3-5 year earlier AAO in carriers of frameshift or structural variants compared to carriers of missense variants.^7^ Furthermore, the group of variants located in the N-terminus of the protein is also associated with an overall earlier AAO.^7^

*PRKN* encodes the Parkin RBR E3 ubiquitin protein ligase. This E3 ubiquitin ligase is recruited to the outer mitochondrial membrane^8^ and activated by phosphorylation of its regulatory Ubiquitin-like (Ubl) domain via PINK1 upon damage-induced mitochondrial membrane depolarization.^9–12^ The disease mechanism is considered to be loss of function.^13–15^ Interestingly, Parkin missing the entire Ubl domain exhibits residual ubiquitination activity.^16,17^ Furthermore, a truncated form of Parkin missing its Ubl domain (Parkin^Δ1–79)^ is translated from an internal translation initiation site (TIS) in Exon 3 at position p.Met80 into the full-length protein (NM_004562.3/NP_004553.2) (Fig. 1).^18–20^ This form is stably detectable, albeit at a low level, in human brain homogenates and cultured cells with or without *PRKN* variants.^19,20^

**Figure 1:**
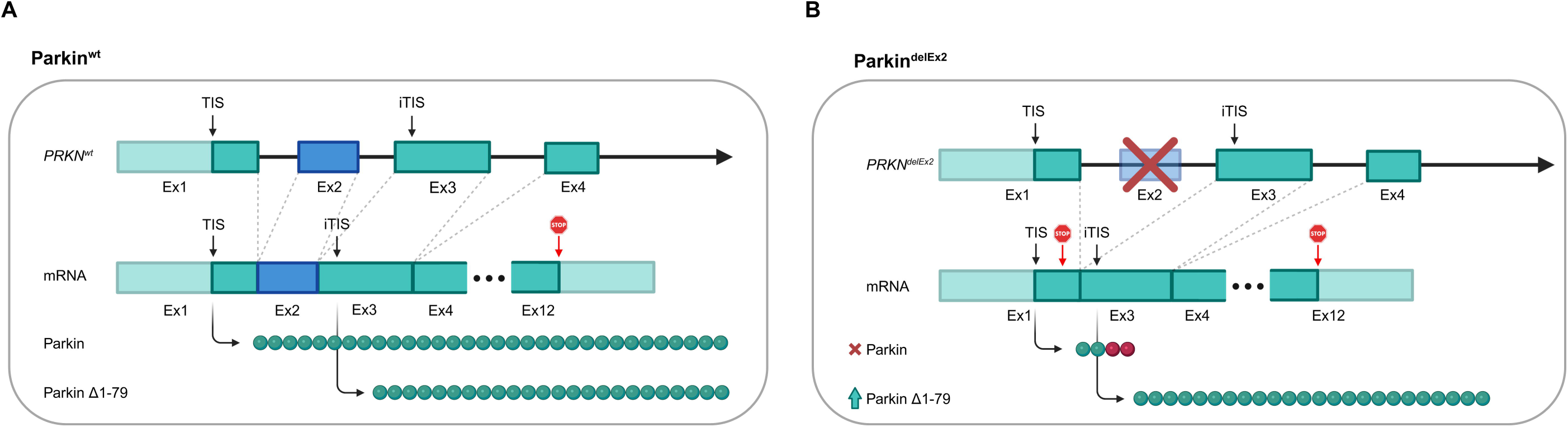
Parkin^Δ1–79^ molecular mechanism. (A) The *PRKN*^wt^ allele translates to an mRNA transcript harboring both the canonical translation initiation site (TIS) and the internal translation initiation site (iTIS) in Exon 3. Thus, in addition to the predominantly expressed canonical full-length Parkin, carriers of wild-type alleles also express the N-terminally truncated Parkin^Δ1–79^ variant, albeit to a lesser degree. (B) The *PRKN*^delEx2^ allele results in a shortened and frameshifted mRNA transcript in which a premature stop codon closely follows the canonical TIS. This likely represents an unfavorable nucleotide context for translation initiation and may shift translation toward the iTIS. We hypothesize a combined effect of TIS favorability shift as well as the closer proximity of the internal translation initiation site to the 5’cap to facilitate increased levels of Parkin^Δ1–79^ and partially remaining function observed in endogenous *in vitro* models of biallelic *PRKN*^delEx2^ carriers. Created with BioRender.

We hypothesized that i) carriers of pathogenic *PRKN* variants located 5’ upstream of this internal TIS might benefit from the compensatory function of Parkin^Δ1–79^ since it will not be affected by these variants, and ii) this might lead to a comparatively milder phenotype with a later AAO. In this study, we indeed demonstrate that carriers of a homozygous Exon 2 deletion in *PRKN* have a later median AAO compared to other PARK-*PRKN* patients. Furthermore, we show elevated Parkin^Δ1–79^ levels with a higher remaining E3 ubiquitin ligase activity compared to carriers of pathogenic variants downstream of the internal TIS using induced dopaminergic midbrain neurons derived from a previously reported asymptomatic homozygous *PRKN*^delEx2^ carrier^21^ and CRISPR/Cas9-edited homozygous *PRKN*^delEx2^ lines derived from healthy controls. Importantly, a recently characterized allosteric Parkin modulator^22,23^ increased the residual but impaired Parkin^Δ1–79^ activity in our models, suggesting a new therapeutic avenue.

## Material and Methods

### Sample and cohort description

To investigate the effect of a biallelic *PRKN*^delEx2^ pathogenic variant on AAO, PARK*-PRKN* patient data from multiple sources was aggregated. Information on biallelic *PRKN* variant carriers curated in the International Parkinson’s Disease and Movement Disorder Society Gene Database (MDSGene) served as the primary data source.^6^ In addition, a recently published PD cohort^24^ and a case report^25^ describing biallelic *PRKN*^delEx2^ variant carriers were included. Furthermore, comprehensive individual data on additional homozygous *PRKN*^delEx2^ variant carriers was collected across multiple research centers through a collaboration with the Department of Neurology, Faculty of Medicine, Juntendo University^26^, the French Parkinson disease Genetics Study Group (PDG)^7^ and the MJFF Global Genetic Parkinson’s Disease Project.^27^ Detailed information, including AAO, age, sex, and country of origin, was collected on a total of 512 PARK*-PRKN* patients. Of those, 26 patients carried a homozygous deletion of Exon 2, and the remaining 485 carried other variants. One of these patients was previously unreported (Table 1). All 26 biallelic *PRKN*^delEx2^ patients were matched to other variant carriers on propensity score (Supplementary Table 1) (see statistics chapter).

**Table 1.** Demographics of patients with a homozygous *PRKN*^delEx2^ variant for *PRKN* (NM_004562.3) and propensity score matched variant carriers included in the AAO analysis.

| ID | Center | Country | Sex | AAO Range | AAE Range | Previously Reported | Previously Included in MDSGene (April 2025) | Match Subclass |
| --- | --- | --- | --- | --- | --- | --- | --- | --- |
| 190 | Istanbul U | Turkey | Male | 31-35 | 36-40 | Yes<sup>7</sup> | No | 17 |
| 792 | Juntendo U | Japan | Male | 21-25 | 51-55 | No | No | 7 |
| 957 | Juntendo U | Japan | Male | 41-45 | 41-45 | Yes<sup>26</sup> | No | 8 |
| 1490 | Juntendo U | Japan | Female | 46-50 | 71-75 | Yes<sup>26</sup> | No | 9 |
| 1610 | Juntendo U | Japan | Male | 41-45 | 51-55 | Yes<sup>26</sup> | No | 10 |
| 1611 | Juntendo U | Japan | Male | 46-50 | 55-60 | Yes<sup>26</sup> | No | 11 |
| 1655 | Juntendo U | Japan | Male | 36-40 | 46-50 | Yes<sup>26</sup> | No | 12 |
| 3248 | Juntendo U | Japan | Female | 31-35 | 51-55 | Yes<sup>26</sup> | No | 13 |
| 4224 | Juntendo U | Japan | Female | 51-55 | 66-70 | Yes<sup>26</sup> | No | 14 |
| 4225 | Juntendo U | Japan | Male | 46-50 | 61-65 | Yes<sup>26</sup> | No | 15 |
| 4662 | Juntendo U | Japan | Female | 31-35 | 51-55 | Yes<sup>26</sup> | No | 16 |
| 761372890 | Sorbonne U | Turkey | Male | 41-45 | NA | Yes<sup>7</sup> | No | 25 |
| VIE-PD3322 | Sorbonne U | Turkmenistan | Female | 11-15 | NA | Yes<sup>7</sup> | No | 26 |
| AR-102 | Central South U | China | Male | 16-20 | NA | Yes<sup>24</sup> | No | 18 |
| AR-168 | Central South U | China | Female | 26-30 | NA | Yes<sup>24</sup> | No | 19 |
| AR-180 | Central South U | China | Female | 21-25 | NA | Yes<sup>24</sup> | No | 20 |
| EOPD-0408 | Central South U | China | Male | 11-15 | NA | Yes<sup>24</sup> | No | 21 |
| EOPD-0615 | Central South U | China | Male | 16-20 | NA | Yes<sup>24</sup> | No | 22 |
| EOPD-0633 | Central South U | China | Female | 26-30 | NA | Yes<sup>24</sup> | No | 23 |
| D81 | Tottori U | Japan | Male | 56-60 | 81-85 | Yes<sup>25</sup> | No | 24 |
| 5 | Barcelona U | Spain | Male | 36-40 | 51-55 | Yes<sup>54</sup> | Yes | 2 |
| 4 | Teikyo U | Japan | Male | 46-50 | 51-55 | Yes<sup>55</sup> | Yes | 5 |
| 7 | IDIBAPS | Spain | Male | 41-45 | 51-55 | Yes<sup>56</sup> | Yes | 1 |
| Patient 2 | Central South U | China | Male | 36-40 | 36-40 | Yes<sup>57</sup> | Yes | 3 |
| 8 | Juntendo U | Japan | Male | 41-45 | 66-70 | Yes<sup>58</sup> | Yes | 6 |
| Cas232 | Pompeu Fabra U | Spain | Male | 51-55 | NA | Yes<sup>59</sup> | Yes | 4 |

AAO = age at onset; AAE = age at examination; U = University; IDIBAPS = Institut d'Investigacions Biomèdiques August Pi i Sunyer.

Using human dermal fibroblasts of one homozygous *PRKN*^delEx2^ carrier, two healthy donors, and four carriers of other homozygous or compound heterozygous *PRKN* variants, we assessed endogenous Parkin activity in vitro (Supplementary Table 2). Additionally, human induced pluripotent stem cells (hiPSC)-derived midbrain dopaminergic neurons (iDN) originating from one homozygous *PRKN*^delEx2^ carrier, two healthy controls, one carrier of a homozygous *PRKN* Exon 7 deletion as well as genome-edited lines derived from the controls carrying homozygous Exon 2 or Exon 3 deletions were analyzed (Supplementary Table 3). The University of Lübeck’s Ethics board has approved the project, as have the local ethics boards from collaborators contributing additional clinical and genetic information on patients not listed in the MDSGene database. All participants gave written informed consent prior to their participation in the study. All previously reported patient IDs were directly adapted from the respective literature and the MDSGene database. Novel IDs were de-identified and are known only to members of the research institutes involved in this study.

### Dopaminergic midbrain neuron differentiation

Differentiation of characterized hiPSCs (Supplementary Fig. 2) into midbrain dopamine neurons was performed following an established protocol with minor alterations (Supplementary Fig. 3).^28,29^ In brief, floorplate induction was initiated in hiPSCs by adding knockout serum replacement (KSR) medium supplemented with 10 µM SB431542 (Tocris) and 100nM LDN-193189 (Stemgent). 100 ng/ml recombinant human Sonic Hedgehog (rhSHH, STEMCELL), 100 ng/ml recombinant human fibroblast growth factor 8a (rhFGF-8a, STEMCELL), and 2 µM Purmorphamine from days 1-5. Further, 3 µM CHIR99021 (STEMCELL) was added from days 3-13. SB431542 and LDN-193189 were withdrawn on days 5 and 11, respectively. KSR medium was gradually phased to Neurobasal medium (NB, gibco) with NeuroCult SM1 neuronal supplement (STEMCELL) (25%, 50%, 75%, 100%) from days 5-10. From days 11-40 neural induction was started by adding 20 ng/ml brain-derived neurotrophic factor (BDNF, STEMCELL), 200 µM ascorbic acid (AA, Sigma), 20 ng/ml glial cell-derived neurotrophic factor (GDNF, STEMCELL), 1 ng/ml transforming growth factor-β3 (TGF-β3, PeproTech), 500 µM cyclic adenosine monophosphate (cAMP, Enzo), and 10 mM DAPT (Tocris). Cells were dissociated with Accutase on day 20 and re-plated on Poly-D-lysine/Laminin/Fibronectin coated 6-well plates as high-density drops of 350,000 cells per 50 µL. From day 40, cells were maintained with NB/SM1. On day 120, dopaminergic midbrain neurons were treated with 1 µM valinomycin in NB/SM1 for 6 h and 14 h or left untreated.

To examine the effects of positive allosteric modulation on Parkin, neurons were pretreated with either 200 µM BIO-2007817 (Probechem) in DMSO (Sigma-Aldrich) or DMSO alone for 1 h followed by 6 h 1 µM valinomycin treatment. Cells were manually scraped and were pelleted by centrifugation at 500 x *g* for 5 min on RT. Pellets were stored at -80 °C until further processing.

### Western blotting and antibodies

For western blots, proteins were isolated from cell pellets by lysis on ice with RIPA buffer supplemented with protease and phosphatase inhibitors. After 30 min incubation on ice, samples were centrifuged at 13,000 x *g* for 20 min. Supernatants were transferred to new tubes, and protein concentration was determined using the DC Protein Assay Kit (Biorad) on a Synergy HT Plate Reader (BioTek). 10 µg protein per sample was loaded and separated on NuPAGE 4-12% Bis-Tris gels (Invitrogen). Proteins were transferred to a nitrocellulose membrane (Protran) and detected with various antibodies: Here, primary antibodies raised against Parkin (Santa Cruz; sc32282; 1:1000), MFN2 (abcam; ab56889; 1:1000), GRP-75 (abcam; ab53098; 1:1 mill), TH (Millipore; AB152; 1:1000), and GAPDH (Cell Signaling; 14C10; 1:50000) were used. Parkin antibody dilution was adjusted for detection in overexpression models of all Parkin constructs (1:1 mill). HRP-conjugated goat anti-mouse (LI-COR; 926-80010; 1:20000) and goat anti-rabbit (LI-COR; 926-80011; 1:20000) were applied as secondary antibodies.

### Image analysis and statistics

Western blots were background-corrected and quantified using Bio-Rad Image Lab (Version 6.1). For background subtraction, the rolling disc method was used. To sensitively assess and normalize ubiquitination efficiency across western blots of *in vitro* models endogenously expressing Parkin via the ratio of ubiquitinated MFN2 (Ub-MFN2) over MFN2, the following formula was applied:

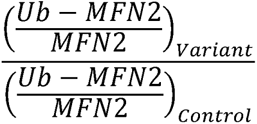

Differences in non-ubiquitinated MFN2 levels were quantified rather than ubiquitinated MFN2 since there is rapid MFN2 turnover and Ub-MFN2 signal reduction in Parkin overexpression models with artificially increased Parkin ubiquitin ligase activity. Data was normalized by the sum of each signal per blot to compare signals without antibody-internal normalization across blots.^30^ To this end, loading schemes were kept consistent across independently conducted western blots for each experiment. All signals were normalized to GAPDH protein levels.

All statistical analyses were performed using R and RStudio. Western blot data was fitted to linear mixed effects models to examine group, treatment time, and interaction effects, including sample ID per independent experiment as the random effect. Models thus had the following general structure:

“lmer(normalizedSignal ∼ Group * TreatmentTime + (1 | ID:Experiment)”

Computation of simple main effects and post-hoc pairwise comparisons using the Kenward-Roger method for degrees of freedom approximation was performed with the emmeans package.^31^ Where linear mixed-effects models failed to pass model assumptions, non-parametric Kruskal-Wallis tests followed by post-hoc Dunn tests or Friedman tests followed by post-hoc Durbin-Conover tests were used instead. Where the assumptions did not hold, and comparisons were limited to two comparisons, Mann-Whitney U tests and Wilcoxon’s tests were applied.

To assess the association of homozygous Exon 2 deletions in *PRKN* with AAO, the data was first propensity score-matched using the covariates sex and country of origin with the MatchIt package.^32^ Subsequently, linear regression models of varying complexities were fitted to the data. To avoid overfitting, the corrected Akaike information criterion (AICc) was computed for each model, selecting the following:

“lm(formula = AAO ∼ delEx2.hmz + Sex + Country, weights = weights)”

The DHARMa package was utilized to diagnose potential model misspecifications and assumption violations.^33^ The marginal hazard ratio and cumulative incidences were calculated using a Cox proportional hazards regression model with clustering by subclassification of the matched data. Cox regression analysis was performed using the survival and adjustedCurves packages.^34,35^ Patients with missing reports on the outcome, predictor, or covariates were excluded from the analysis. All post-hoc tests were Holm corrected. All tests were two-tailed, and the alpha threshold probability was set to 0.05.

## Results

### Biallelic *PRKN*^delEx2^ variant carriers develop Parkinson’s disease at a later age compared to PARK-*PRKN* patients carrying other variants

A review of the MDSGene database revealed that a homozygous *PRKN*^delEx2^ variant causes PD with a later AAO compared to carriers of other *PRKN* variants as indicated by a higher median AAO (39.5 vs. 31 years) and the identification of an individual in their 60s, unaffected at the time of clinical examination.^21^ To explore this further, we reached out to the principal investigators of several larger patient cohorts and analyzed 26 *PRKN*^delEx2^ carriers and 26 carriers of other *PRKN* variants with detailed demographic and clinical data, who were matched on the propensity score, including covariates such as age, sex, and country of origin. We fitted a linear regression model to predict AAO, using a binary indicator variable for homozygous *PRKN*^delEx2^ variant status with sex and country of sample origin as covariates. Our model indicates an association between homozygous variant status and AAO (Δμ[= 13.22, t(45) = 4.88, p < 0.001) (Fig. 2A). Furthermore, we computed the marginal hazard ratio and cumulative incidence for biallelic *PRKN*^delEx2^ variant carriers compared to matched variant carriers using a Cox proportional hazards model, estimating a hazard ratio reduction of 64% in biallelic *PRKN*^delEx2^ carriers (HR = 0.36, z = - 3.95, p < 0.001) (Fig. 2B).

**Figure 2:**
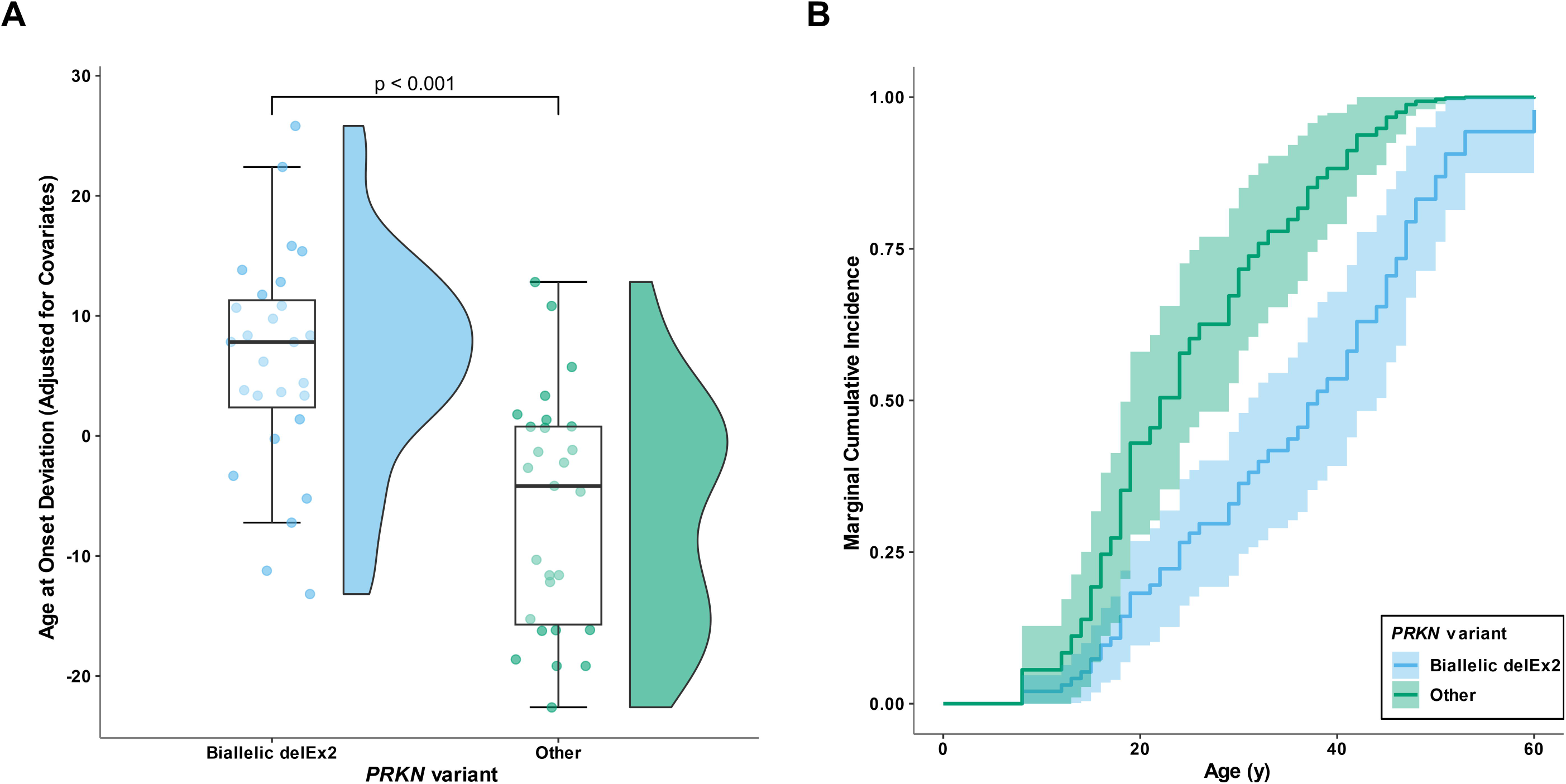
Later age at onset in biallelic *PRKN*^delEx2^ carriers. (A) Partial residual plot highlighting the impact of a biallelic *PRKN*^delEx2^ variant on age at onset (AAO) compared to matched carriers of other pathogenic *PRKN* variants with an estimated average marginal effect of 13.22 years (t(45) = 4.88, p < 0.001). Covariates corrected for in the underlying linear regression model were sex and country of origin. (B) A Cox proportional hazards model to estimate the marginal cumulative incidence and hazard ratio for biallelic *PRKN*^delEx2^ variant carriers and matched variant carriers (HR = 0.36 SE = 0.26, z = - 3.79, p < 0.001). Sample size: n = 26 per group. The significance threshold was set to p = 0.05. Reported standard errors are cluster robust.

Additionally, we classified all patients more generally by variant, located either upstream or downstream of the internal TIS, starting at position c.238, and thus for its potential of translating the Parkin^Δ1–79^ protein (see below). *PRKN*^delEx1^ variants were not classified as upstream due to promoter involvement and loss of the canonical translation start site. Associations with AAO of carrying one or two alleles with variants upstream of the internal TIS were determined by extension of the *PRKN*^delEx2^ multilinear regression model, and thus without prior propensity score matching, as the matched data for these additional binary indicators did not pass model assumptions. In contrast to a biallelic *PRKN*^delEx2^ variant, the presence of at least one variant (Δμ[= -1.98, t(471) = -1.23, p = 0.216) or variants exclusively upstream of the internal TIS (Δμ[= -0.74, t(471) = -0.45, p = 0.654) was not associated with AAO when compared to patients only carrying variants downstream of the internal TIS. Similarly, in a total of 20 compound heterozygous patients (Supplementary Table 4), a single *PRKN*^delEx2^ allele was not associated with AAO (Δμ[= -1.74, t(471) = -0.65, p = 0.515). Further stratifying the patients by variant impact, where missense variants were classified as ‘moderate impact’ and all other variants as ‘high impact’, did not improve model fit and did not reveal an association of variant impact with AAO or interaction effects with the variant position binary indicator.

### *In vitro* overexpression of *PRKN*^delEx2^ retains partially functional Parkin**^Δ^**^1–79^

Previous studies demonstrated the expression of a truncated Parkin isoform (Parkin^Δ1–79)^ lacking the Ubl domain from an internal TIS at position c.238 of the wildtype transcript.^19,20^ Furthermore, artificial constructs lacking the Ubl domain, similar to Parkin^Δ1–79^, retained partial function as measured by substrate ubiquitination, mitophagy, and mitochondrial recruitment.^16,17,36^ We hypothesized that the later AAO observed in carriers of the *PRKN*^delEx2^ may be due to the expression of Parkin^Δ1–79^ with its residual function. Notably, the deletion of Exon 2 in *PRKN* does not involve the internal TIS located in Exon 3 and shifts this alternative TIS closer to the 5’regulatory region of the transcript due to the deletion (Fig. 1B).

First, we investigated Parkin levels and its E3 ubiquitin ligase activity in an *in vitro* overexpression model following mitochondrial depolarization periods using valinomycin (Fig. 3). For this, neuroblastoma (SH-SY5Y) cells with a knockout of endogenous *PRKN* (*PRKN* KO due to a genome-edited Exon 3 deletion) were used to overexpress either the *PRKN*^delEx2^ variant, a truncated *PRKN* variant starting at the internal TIS (*PRKN*^c.1-237del^), or wild-type *PRKN* (*PRKN*^wt^). We analyzed Parkin^Δ1–79^ levels in these lines (Fig. 3A-D). Western blotting showed comparable levels of Parkin^Δ1–79^ for all PRKN-containing constructs in non-treated cells and after 3 h treatment with valinomycin. However, at the 14 h-timepoint of treatment, levels of Parkin^Δ1–79^ increased in *PRKN*^c.1-237del^ transduced cells (F(2,13.48) = 8.65, p = 0.004) both compared to *PRKN*^wt^ (Δμ = 15.16, t(13.48) = 3.21, p = 0.013) and to *PRKN*^delEx2^ (Δμ = 18.44, t(13.48) = 3.90, p = 0.005) (Fig. 3C). This is furthermore reflected by a change of Parkin^Δ1–79^ for *PRKN*^c.1-237del^ cells over increased mitochondrial depolarization times (F(2,12) = 4.08, p = 0.045), where levels increased between 3 h and 14 h of depolarization (Δμ = 10.20, t(12) = 2.81, p = 0.048) (Fig. 3D). Total Parkin expression (full-length Parkin + Parkin^Δ1–79)^ was 41%-62% reduced in *PRKN*^delEx2^ and *PRKN*^c.1-237del^ cells compared to neuroblastoma cells overexpressing *PRKN*^wt^ across all treatment time points (Supplementary Fig. 4A).

**Figure 3:**
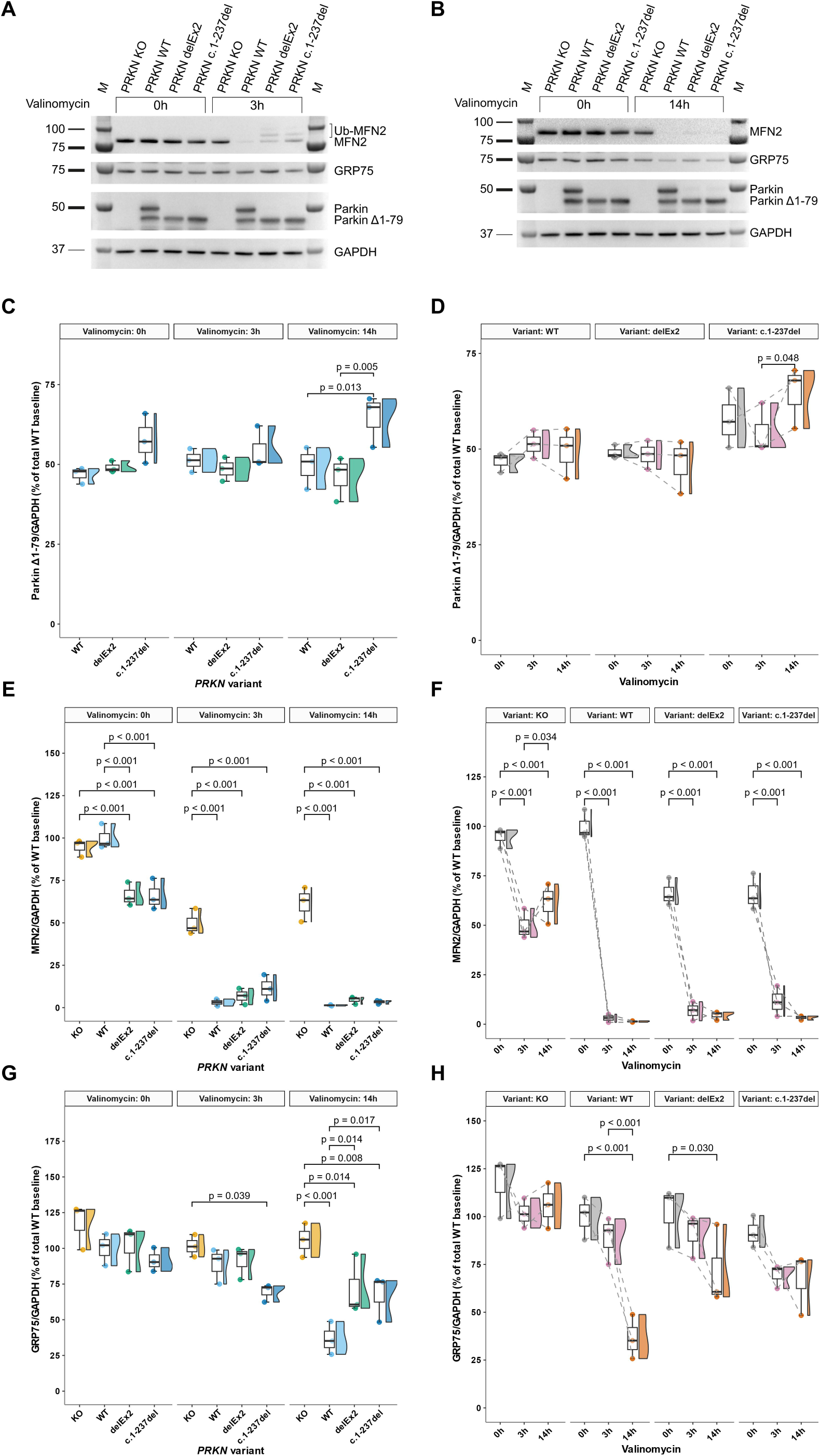
Overexpression of three *PRKN* constructs in a *PRKN* knockout neuroblastoma cell model highlights Parkin^Δ1–79^ activity. (A-B) Western blot analysis of MFN2, GRP75, and Parkin in *PRKN*-KO SH-SY5Y neuroblastoma cells overexpressing *PRKN*^wt^, *PRKN*^delEx2^, or *PRKN*^c.1-237del^, a shortened construct, starting directly at the internal translation initiation site. Mitochondrial depolarization via 1 μM valinomycin was performed for 3 h (A) and 14 h (B), respectively. Signals were normalized against GAPDH. (C-D) Parkin^Δ1–79^ expression was comparable between all overexpression models (C) and remained stable after prolonged valinomycin treatment (D). (E-F) Between- (E) and within-group (F) differences in MFN2 expression. (G-H) Differences in GRP75 levels between groups (G) and over time (H). Sample size: n = 3 from independently repeated experiments across three cell passages. The significance threshold was set to p = 0.05. Pairwise comparisons of linear mixed effects model derived estimated marginal means (C-H) were Holm-adjusted.

To assess Parkin ubiquitin ligase activity, changes in levels of MFN2, a direct substrate of Parkin, were quantified (Fig. 3 A-B & E-F). MFN2 levels were reduced in both non-treated *PRKN*^delEx2^ and *PRKN*^c.1-237del^ cells compared to *PRKN*^KO^ or *PRKN*^wt^ cells (F(3,24) = -25.01, p < 0.001), (Δμ < -28.34, t(24) < -5.54, p < 0.001) (Fig. 3E). Following valinomycin treatment, MFN2 levels dropped in all overexpression models (F(2, 16) < -88.82, p < 0.001), but also in the non-transduced cells (F(2, 16) = -41.26, p < 0.001) (Fig. 3F). However, the MFN2 level difference between the non-transduced cells and all overexpression models increased with prolonged mitochondrial depolarization after 3 h (Δμ < -38.28, t(24) < -7.48, p < 0.001) and again after 14 h (Δμ < -57.08, t(24) < -11.15, p < 0.001). Moreover, MFN2 levels partially recovered in the non-transduced cells comparing 3 h and prolonged 14 h valinomycin treatments (Δμ = 11.83, t(16) = 2.31, p = 0.034).

To further assess mitochondrial dynamics and turnover, protein levels of the mitochondrial matrix protein GRP75 were determined (Fig. 3G-H). Compared to the non-transduced cells, GRP75 levels were significantly reduced in *PRKN*^c.1-237del^ cells upon 3 h valinomycin treatment (Δμ = -32.10, t(23.76) = 2.98, p = 0.039) and reduced in all Parkin overexpression models following 14 h mitochondrial depolarization (F(3,23.76) = -13.86, p < 0.001), (Δμ < - 34.26, t(23.76) < -3.18, p < 0.014) (Fig. 3G). Upon prolonged valinomycin treatment, GRP75 levels decreased in *PRKN*^wt^ (F(2,16) = -21.32, p < 0.001) and *PRKN*^delEx2^ cells (F(2,16) = - 4.38, p = 0.030), with a borderline significant reduction in *PRKN*^c.1-237del^ cells (F(2,16) = - 3.35, p = 0.061).

### Parkin**^Δ^**^1–79^ is expressed in SH-SY5Y cells with a frameshift variant upstream of the internal TIS and displays residual mitochondrial translocation

We hypothesize that, besides the specific case of a *PRKN*^delEx2^ variant, any truncating *PRKN* variant located upstream of the internal TIS that does not affect cis-regulatory elements of *PRKN* should not negatively affect the level of Parkin^Δ1–79^ and, given a shift in TIS favorability, could even alter its abundance.

We therefore analyzed levels of endogenous Parkin protein as total (combined full-length Parkin and Parkin^Δ1–79)^ and Parkin^Δ1–79^ separately upon valinomycin-induced mitochondrial depolarization by western blot in wild-type, *PRKN*^c.100_101insC^, and *PINK1*-KO SH-SY5Y cell lines. In *PINK1*-KO cells, in contrast to wild-type cells, levels of both Parkin forms did not decrease upon prolonged treatment with valinomycin (Fig. 4A-D, Supplementary Fig. 5A- D).^37^ Instead, with prolonged mitochondrial depolarization, levels of total Parkin (F(3,18) = 13.35, p < 0.001) and Parkin^Δ1–79^ (F(3,18) = 7.45, p = 0.002) increased by up to 80% (Supplementary Fig. 5B) and 62% (Fig. 4D) respectively. In contrast, Parkin^Δ1–79^ levels and total Parkin in wild-type cells remained at lower levels (Δμ < -20.48, t(20.7) < -3.07, p < 0.018) (Fig. 4C), (Δμ < -32.82, t(23.8) < -2.80, p < 0.030) (Supplementary Fig. 5A), and comparatively constant (F(3,18) = 0.91, p = 0.457) (Fig. 4D), (F(3,18) = 1.00, p = 0.414) (Supplementary Fig. 5B) following valinomycin treatment. As expected, no full-length Parkin was detected in *PRKN*^c.100_101insC^ cells (Fig. 4A-B). Similarly to wild-type cells, we only observed a borderline significant increase in Parkin^Δ1–79^ levels over time in *PRKN*^c.100_101insC^ cells (F(3,18) = 2.97, p = 0.060) (Fig. 4D) at significantly lower levels compared to the *PINK1*-KO after 3 h and 14 h of mitochondrial depolarization (Δμ < -18.01, t(20.7) < -2.70, p < 0.041) (Fig. 4C). Levels of full-length Parkin gradually decreased with prolonged mitochondrial depolarization in wild-type SH-SY5Y cells (F(3,16) = 3.42, p = 0.043), indicating continuous autoubiquitination of full-length Parkin (Supplementary Fig. 5D). In contrast to our findings in *PRKN*^delEx2^ variant carriers (Fig. 5E), Parkin^Δ1–79^ levels were not significantly increased in *PRKN*^c.100_101insC^ cells compared to wild-type cells (Fig. 4D).

**Figure 4:**
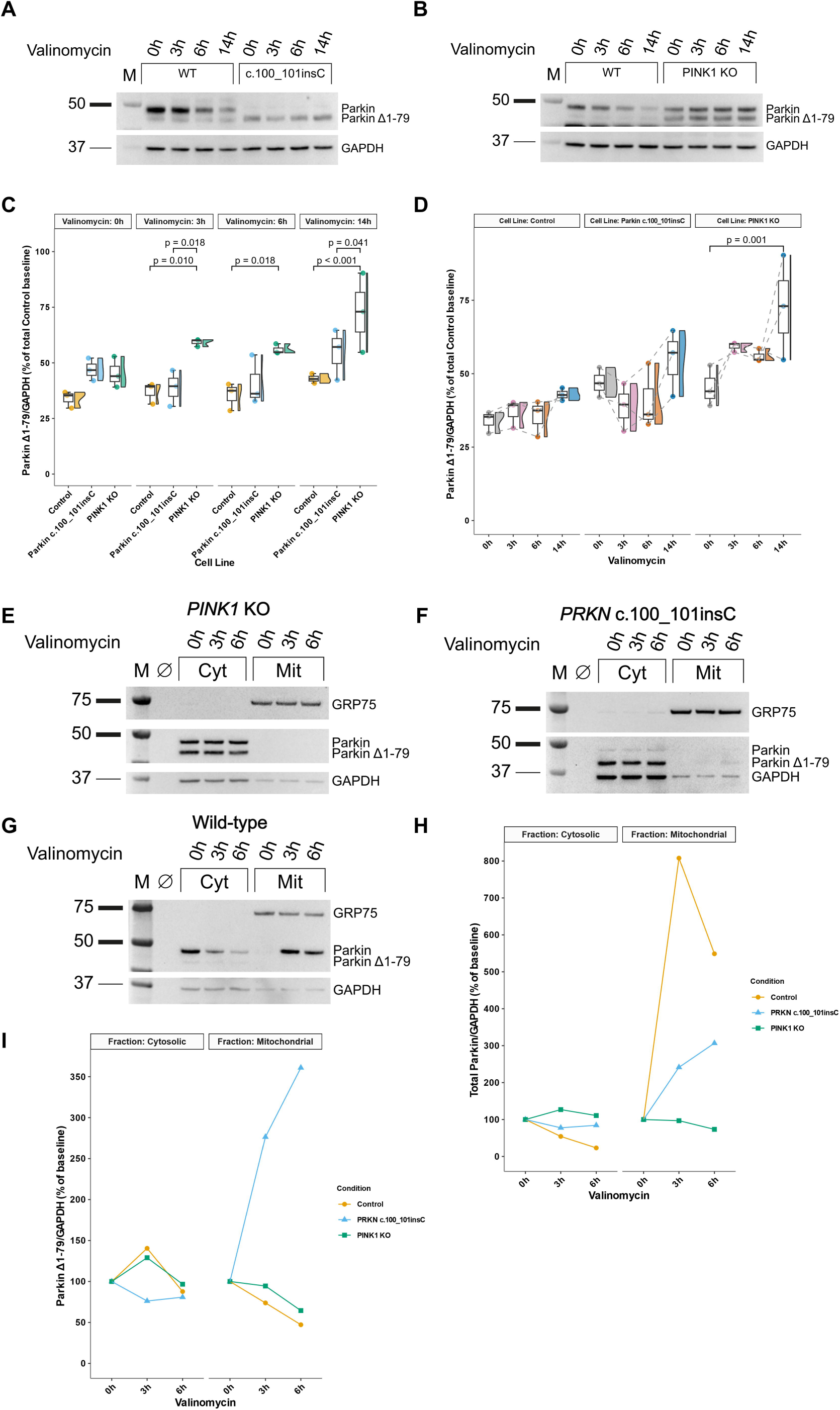
Endogenous Parkin expression and subcellular localization in wild-type, *PRKN*^c.100_101insC^, and *PINK1* knockout neuroblastoma cells. (A-B) Western blot analysis of endogenous Parkin in wild-type, *PRKN*^c.100_101insC^ (A), and *PINK1*-KO (B) SH-SY5Y neuroblastoma cells following mitochondrial depolarization with 1 μM valinomycin across multiple time points. The c.100_101insC *PRKN* variant introduces a frameshift in Exon 2 of *PRKN*, resulting in premature termination of full-length Parkin within five amino acids of the duplication site. Expression of Parkin^Δ1–79^, initiated upstream of the frameshift variant, remains intact. Signals were normalized against GAPDH. (C-D) Parkin^Δ1–79^ protein levels between groups (C) and over time (D). (E-G) Assessment of Parkin translocation after 1 μM valinomycin-induced mitochondrial depolarization by western blot analysis. Cytosolic and mitochondrial fractions were isolated from *PINK1*-KO (E), *PRKN*^c.100_101insC^ (F), and wild-type (G) neuroblastoma cells. (H-I) Deviations from the cytosolic and mitochondrial baseline of total Parkin (H) and Parkin^Δ1–79^ levels (I). Sample size: n = 3 from independently repeated experiments across three cell passages (A-D) and n = 1 (E-I). The significance threshold was set to p = 0.05. Pairwise comparisons of linear mixed effects model derived estimated marginal means (C-D) were Holm-adjusted.

**Figure 5:**
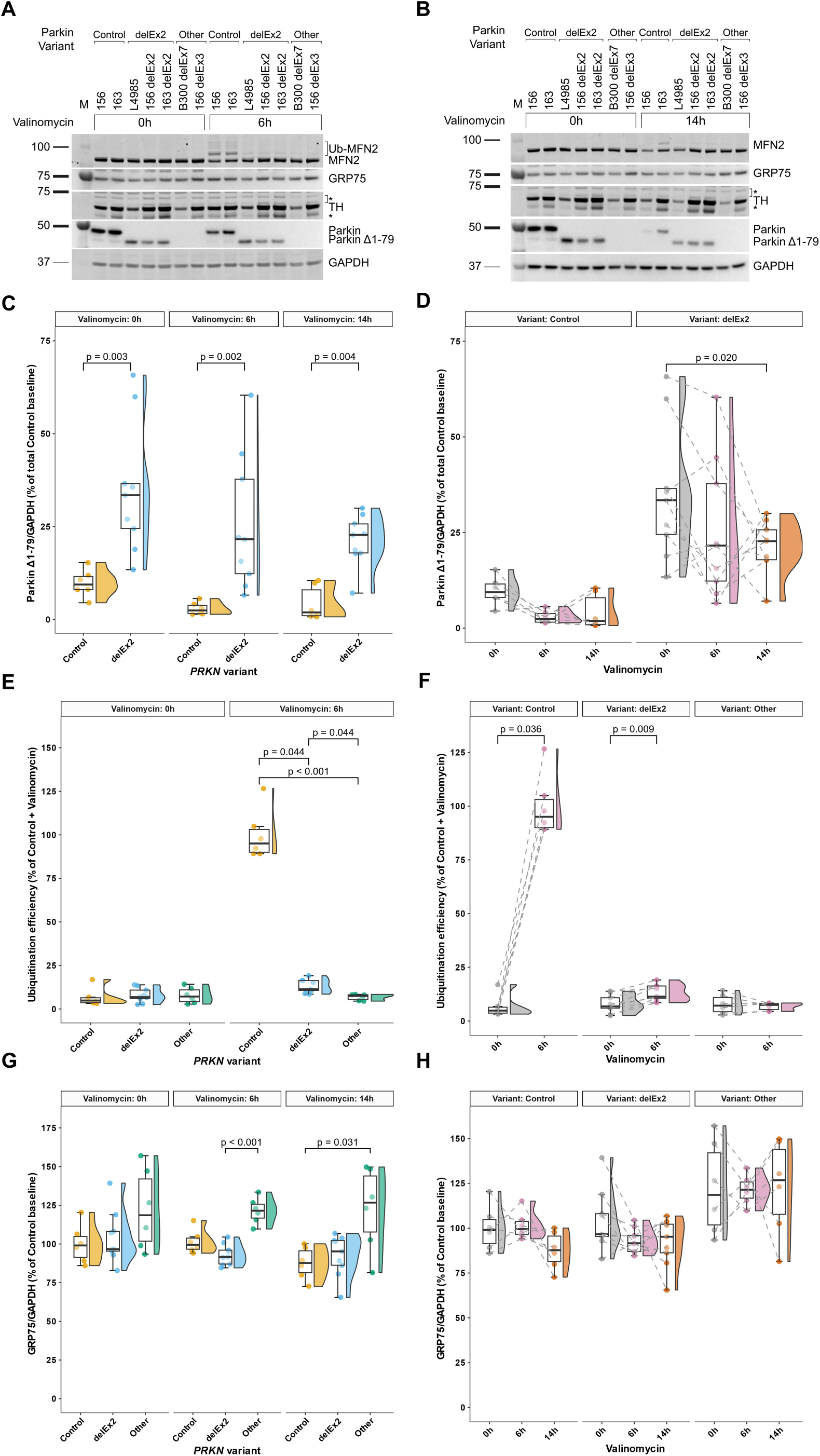
Residual endogenous Parkin function and increased Parkin^Δ1–79^ levels in biallelic *PRKN*^delEx2^ patient-derived dopaminergic midbrain neurons. (A-B) Western blot analysis of MFN2, GRP75, TH, and Parkin in hiPSC-derived midbrain dopaminergic neurons of healthy controls (n = 6), iso- and heterogenic biallelic *PRKN*^delEx2^ carriers (n = 9), as well as carriers of other biallelic *PRKN* exon deletions (n = 6) following 1 μM valinomycin treatment for 6 h (A) and 14 h (B). Signals were normalized against GAPDH. (C-D) Parkin^Δ1–79^ expression differences between groups (C) and expression changes with increased mitochondrial depolarization periods (D). (E-F) Differences in MFN2 ubiquitination between *PRKN* variant carriers (E) and MFN2 ubiquitination change over time (F). (G-H) Comparison of GRP75 expression between *PRKN* variant carriers (G) and over prolonged valinomycin treatment (H). Sample sizes represent a combination of individual patient-derived neurons and independently repeated experiments across three differentiations. The significance threshold was set to p = 0.05. Post-hoc multiple comparisons via Dunn tests (E, G) and Durbin-Conover tests (D) were Holm-adjusted.

To investigate whether previous findings of residual mitochondrial translocation of overexpressed ΔUbl-Parkin variants^10,16,38^ could be replicated for endogenous Parkin^Δ1–79^, we quantified Parkin in cytosolic and mitochondrial cellular fractions following mitochondrial depolarization in all three SH-SY5Y cell lines (Fig. 4E-I). As expected, no mitochondrial translocation of Parkin was observed in the *PINK1*-KO (Fig. 4E). Notably, in *PRKN*^c.100_101insC^ cells (Fig. 4F, I), but not in wild-type cells (Fig. 4G, I), levels of mitochondrial Parkin^Δ1–79^ increased by 260% after 6 h valinomycin treatment when compared to the non-treated cells. Furthermore, total mitochondrial Parkin levels increased by 700% in wild-type cells at the 3 h-time point but subsequently decreased by 250% after mitochondrial depolarization for 6 h (Fig. 4G, H).

### Residual endogenous Parkin function in *PRKN*^delEx2^ patient-derived midbrain dopaminergic neurons

To assess whether the effects seen in the neuroblastoma models of Parkin^Δ1–79^ would also translate to more relevant, patient-derived endogenous cell models, we analyzed three independent differentiations of iPSC-derived dopaminergic midbrain neurons (iDN) from two PARK*-PRKN* patients carrying a homozygous deletion of either Exon 2 or Exon 7, three genome-edited isogenic cells carrying a biallelic Exon 2 deletion or a biallelic Exon 3 deletion, and two healthy controls.

The cells were analyzed under basal conditions and upon valinomycin-induced mitochondrial depolarization. In biallelic *PRKN*^delEx2^ carriers, no full-length Parkin could be detected by western blot analysis (Fig. 5A-B), but increased Parkin^Δ1–79^ levels (r < -0.93, W < 2, p < 0.004) (Fig. 5C) were present when compared to controls. Parkin^Δ1–79^ levels decreased with prolonged mitochondrial depolarization in carriers of biallelic *PRKN*^delEx2^ (W_Kendall_ = 0.38, χ2(2) = 6.89, p = 0.032) with a similar, albeit non-significant trend in controls (W_Kendall_ = 0.33, χ2(2) = 4.00, p = 0.135) (Fig. 5D). While total Parkin was higher under basal conditions in healthy controls (r = 0.82, W = 49, p = 0.011), Parkin^Δ1–79^ was more stable in *PRKN*^delEx2^ carriers after 14 h of mitochondrial depolarization (r = -0.70, W = 8, p = 0.029, n_obs_ = 15) (Supplementary Fig. 6A-B). As expected, in iDN derived from either the isogenic Exon 3 deletion line or the patient-derived homozygous Exon 7 deletion line, we detected neither full-length Parkin nor Parkin^Δ1–79^. Importantly, *PRKN*^delEx2^ patients displayed *PRKN* mRNA levels comparable to healthy controls, whereas transcript levels in carriers of other exon deletions were non-significantly reduced by about 40-50% on average (Δμ < -38.83, t(4) > -2.75, p > 0.155) (Supplementary Fig. 6C).

To assess endogenous Parkin ubiquitin ligase activity in *PRKN*^delEx2^ carriers, we measured the ubiquitination efficiency by calculating the ratio between ubiquitinated MFN2 and non-ubiquitinated MFN2 after 6 h of mitochondrial depolarization in iDN (Fig. 5A-D), but also in human dermal fibroblast cultures (Supplementary Fig. 7A-D). As expected, iDN derived from healthy controls exhibited significantly higher Parkin ubiquitination efficiency compared to both *PRKN*^delEx2^ neurons and neurons carrying other exon deletions (r > 0.59, z > 2.29, p < 0.044). Importantly, biallelic *PRKN*^delEx2^ carriers displayed significantly higher ubiquitination efficiency compared to carriers of other variants (r = 0.59, z = 2.29, p = 0.044) (Fig. 5E). The within-group increase after mitochondrial depolarization further supported this observation (r = -1, V = 0, p = 0.009) (Fig. 5F). Analogous experiments in human dermal fibroblast cultures replicated these findings (Supplementary Fig. 7A-D).

Quantification of the mitochondrial matrix protein GRP75 indicated a reduction in GRP75 levels for both *PRKN*^delEx2^ carriers after 6 h (r = 0.97, z = 3.77, p < 0.001) and healthy controls after 14 h (r = 0.74, z = 2.56, p = 0.031) of valinomycin treatment when compared to carriers of other *PRKN* variants (Fig. 5G). We did not detect significant changes in GRP75 levels upon prolonged valinomycin treatment within a specific variant group (W_Kendall_ < 0.23, χ2(2) < 4.22, p > 0.121) (Fig. 5H).

### Molecular glue BIO-2007817 increases endogenous Parkin**^Δ^**^1–79^ activity

Recently, tetrahydropyrazolo-pyrazine (THPP) compounds have been shown to act as a molecular glue between the RING0 domain of Parkin and phosphorylated ubiquitin, thereby promoting a secondary, Ubl-independent Parkin activation mechanism.^17,22, 23^ Based on the partial rescue of similar ΔUbl variants achieved with THPP treatment,^22^ we hypothesized its possible beneficial effects on endogenous Parkin^Δ1–79^, expressed by *PRKN*^delEx2^ carriers.

Consequently, we treated iDNs with the THPP compound BIO-2007817 and determined differences in ubiquitination efficiency by measuring ubiquitination of MFN2 as well as Parkin levels after 6 h of mitochondrial depolarization by western blot (Fig. 6A). In general, ubiquitination efficiency was higher in healthy controls and *PRKN*^delEx2^ carriers compared to carriers of other *PRKN* exon deletions (Δμ > 16.06, t(9.14) > 4.15, p < 0.002) in both, THPP-treated and untreated cells. Furthermore, ubiquitination efficiency was significantly higher in healthy controls compared to *PRKN*^delEx2^ carriers with and without treatment (Δμ > 28.86, t(9.14) > 8.45, p < 0.001) (Fig. 6B). Importantly, ubiquitination efficiency increased by 30% in *PRKN*^delEx2^ carriers treated with THPP compared to DMSO-treated cells (Δμ = 7.26, t(9.14) = 3.46, p = 0.014), or to 31% ubiquitination efficiency relative to the DMSO-treated healthy controls (Fig. 6C). Notably, THPP treatment induced a 40% ubiquitination efficiency reduction in healthy controls (Δμ = -39.96, t(6) = 16.49, p < 0.001).

**Figure 6:**
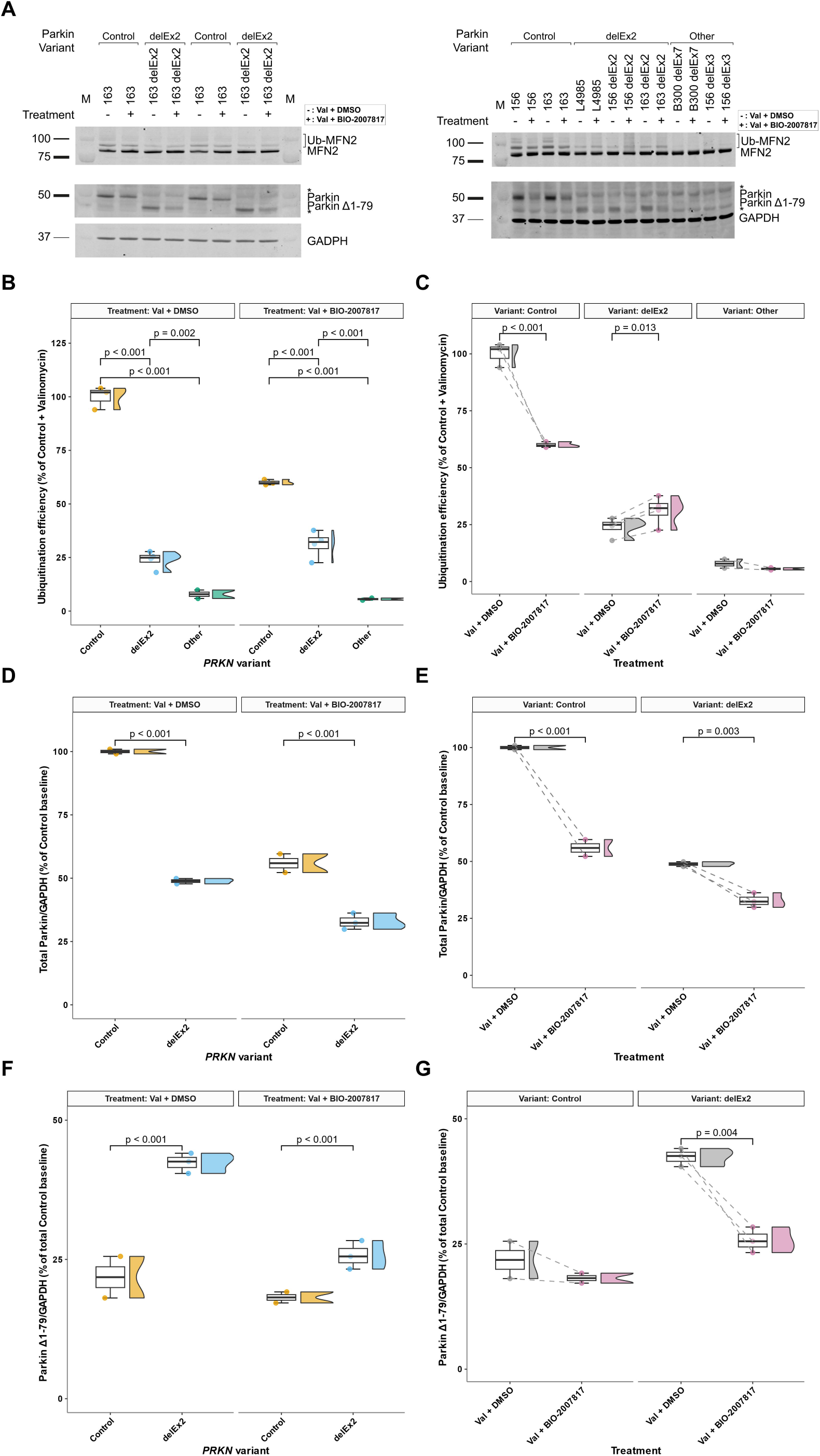
BIO-2007817 partially rescues endogenous Parkin function in biallelic *PRKN*^delEx2^ patient-derived dopaminergic midbrain neurons. (A) Western blot analysis of MFN2 and Parkin in iPSC-derived dopaminergic midbrain neurons of healthy controls (n = 3), carriers of biallelic *PRKN*^delEx2^ (n = 4), or other biallelic exon deletion variants (n = 2) following treatment with an allosteric Parkin activator. All neurons were preincubated for 1 h with either DMSO or 200 μM BIO-2007817, followed by adding 1 μM valinomycin for a further 6 h. Signals were normalized against GAPDH. (B) Between-group differences of MFN2 ubiquitination efficiency with and without BIO-2007817 treatment. (C) Effect of BIO-2007817 on MFN2 ubiquitination efficiency per *PRKN* variant group. (D-E) Total Parkin (full-length Parkin + Parkin^Δ1–79^) protein level differences between groups (D) and its BIO-2007817 induced change (E). (F-G) Variation of Parkin^Δ1–79^ expression between (F) and within groups (G). As cross-blot normalization was not feasible for the analysis of Parkin levels here, only the right blot (A) was included in the analysis. Sample sizes represent a combination of individual patient-derived neurons and independently repeated experiments across two differentiations. The significance threshold was set to p = 0.05. Pairwise comparisons of linear mixed effects model derived estimated marginal means (C-F) were Holm-adjusted.

Levels of total Parkin were higher in healthy controls compared to *PRKN*^delEx2^ carriers in both conditions (Δμ > 23.09, t(5.12) > 8.60, p < 0.001) (Fig. 6D) and decreased with THPP treatment in both groups (Δμ < -16.02, t(3) < -3.46, p < 0.003) (Fig. 6E). In contrast, in DMSO-treated cells, levels of Parkin^Δ1–79^ were overall higher in *PRKN*^delEx2^ carriers compared to healthy controls (Δμ > 7.55, t(5.84) > 2.88, p < 0.029) (Fig. 6F), comparable to our previous findings (Fig. 5E). With THPP treatment, Parkin^Δ1–79^ significantly decreased in *PRKN*^delEx2^ (Δμ = -16.62, t(3) = -7.76, p = 0.004), but not in healthy control neurons (Δμ = - 3.63, t(3) = -1.39, p = 0.260) (Fig. 6G).

### *In silico* translation initiation prediction for Parkin**^Δ^**^1–79^

Recently, *in silico* tools based on deep learning models have been developed to determine, score, and annotate translation initiation sites.^39–42^ Hence, we scored three different *PRKN* transcripts for the potential expression of Parkin^Δ1–79^: *PRKN*^wt^, *PRKN*^delEx2^, as well as the *PRKN*^c.100_101insC^ (p.Q34Pfs*5) frameshift variant using the TIS Transformer tool (Table 2).^42^ As expected, for the full-length, wild-type transcript of *PRKN*, the canonical TIS was predicted with the highest probability score at 0.94 and the internal TIS at position c.238 scored <0.01. In contrast, analysis of the *PRKN*^delEx2^ transcript scored the internal TIS highest (0.96). For *PRKN*^c.100_101insC^, the canonical TIS probability was 0.11, and an out-of-frame TIS at position c.32 was 0.57. The internal TIS in *PRKN*^c.100_101insC^ was scored low as in wild-type *PRKN* (<0.1). Notably, the reading frame would move back in-frame at position c.100 for this transcript due to the frameshift variant. This should result in the Parkin isoform that is 2.5 kDa smaller than the full-length Parkin and differs from the wild-type Parkin in 25 amino acids at the N terminus. While this difference most likely leads to protein instability, a faint band of appropriate size observed in some western blots of *PRKN*^c.100_101insC^ cells may match this prediction (Fig. 4 F). In this context, rendering the highest scoring out-of-frame TIS non-functional for the c.100_101insC variant by introducing an additional missense variant (c.33T>A) increased the internal TIS prediction score to 0.86. Importantly, with a single missense variant in the canonical TIS (c.2T>C) TIS Transformer called the internal TIS at a probability of 0.89.

**Table 2.**
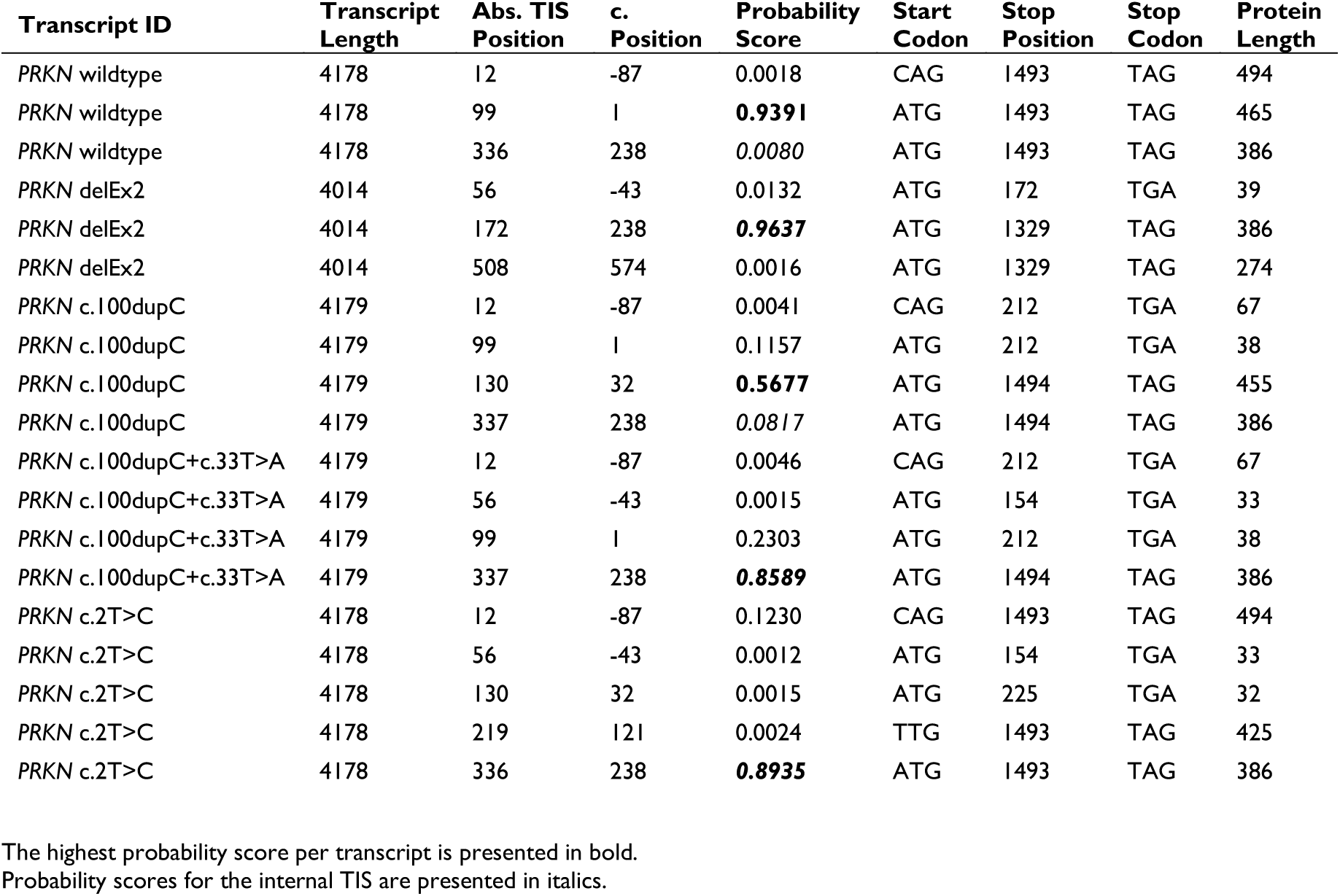
*In silico* TIS prediction by TIS Transformer.

| Transcript ID | Transcript Length | Abs. TIS Position | c. Position | Probability Score | Start Codon | Stop Position | Stop Codon | Protein Length |
| --- | --- | --- | --- | --- | --- | --- | --- | --- |
| *PRKN* wildtype | 4178 | 12 | -87 | 0.0018 | CAG | 1493 | TAG | 494 |
| *PRKN* wildtype | 4178 | 99 | 1 | **0.9391** | ATG | 1493 | TAG | 465 |
| *PRKN* wildtype | 4178 | 336 | 238 | *0.0080* | ATG | 1493 | TAG | 386 |
| *PRKN* delEx2 | 4014 | 56 | -43 | 0.0132 | ATG | 172 | TGA | 39 |
| *PRKN* delEx2 | 4014 | 172 | 238 | **0.9637** | ATG | 1329 | TAG | 386 |
| *PRKN* delEx2 | 4014 | 508 | 574 | 0.0016 | ATG | 1329 | TAG | 274 |
| *PRKN* c.100dupC | 4179 | 12 | -87 | 0.0041 | CAG | 212 | TGA | 67 |
| *PRKN* c.100dupC | 4179 | 99 | 1 | 0.1157 | ATG | 212 | TGA | 38 |
| *PRKN* c.100dupC | 4179 | 130 | 32 | **0.5677** | ATG | 1494 | TAG | 455 |
| *PRKN* c.100dupC | 4179 | 337 | 238 | *0.0817* | ATG | 1494 | TAG | 386 |
| *PRKN* c.100dupC+c.33T>A | 4179 | 12 | -87 | 0.0046 | CAG | 212 | TGA | 67 |
| *PRKN* c.100dupC+c.33T>A | 4179 | 56 | -43 | 0.0015 | ATG | 154 | TGA | 33 |
| *PRKN* c.100dupC+c.33T>A | 4179 | 99 | 1 | 0.2303 | ATG | 212 | TGA | 38 |
| *PRKN* c.100dupC+c.33T>A | 4179 | 337 | 238 | **0.8589** | ATG | 1494 | TAG | 386 |
| *PRKN* c.2T>C | 4178 | 12 | -87 | 0.1230 | CAG | 1493 | TAG | 494 |
| *PRKN* c.2T>C | 4178 | 56 | -43 | 0.0012 | ATG | 154 | TGA | 33 |
| *PRKN* c.2T>C | 4178 | 130 | 32 | 0.0015 | ATG | 225 | TGA | 32 |
| *PRKN* c.2T>C | 4178 | 219 | 121 | 0.0024 | TTG | 1493 | TAG | 425 |
| *PRKN* c.2T>C | 4178 | 336 | 238 | **0.8935** | ATG | 1493 | TAG | 386 |

The highest probability score per transcript is presented in bold. Probability scores for the internal TIS are presented in italics.

## Discussion

With this translational study, we propose a link between the endogenous residual function of truncated Parkin^Δ1–79^, present at elevated levels in biallelic *PRKN*^delEx2^ variant carriers, and a later AAO of PD in such carriers compared to that in carriers of other *PRKN* pathogenic variants. Promisingly, we provide first evidence for allosteric modulation of endogenously expressed Parkin^Δ1–79^ in a patient-derived *in vitro* PD model by a THPP compound that partially restored ubiquitination activity. Thus, these findings have three important implications: i) a *PRKN*^delEx2^ variant-specific later PARK*-PRKN* disease onset compared to carriers of other variants, ii) opportunities for future personalized counseling and treatment strategies in patients with residual Parkin^Δ1–79^ expression, and iii) insights for *PRKN*-variant research and successful gene knockout design taking into account residual functionality.

Using a combination of MDSGene database entries^6^, recently published cohorts,^24,25^ and additional patient data collected in a multi-site collaborative effort, we investigated whether elevated Parkin^Δ1–79^ expression *in vitro*, particularly in biallelic carriers of *PRKN*^delEx2^ variants, would translate to a specific clinical manifestation of PD by affecting the median AAO. Our linear model on propensity score matched PARK*-PRKN* patients estimated a later AAO in biallelic *PRKN*^delEx2^ variant carriers compared to carriers of other variants by 13.22 ± 2.71 years on average, likely linked to increased Parkin^Δ1–79^ levels and its partially remaining function. Of note, biallelic *PRKN*^delEx2^ carriers also displayed a wide AAO range (13-60 years). This wide range warrants further research, investigating potential causes for this variation and whether it is linked to differences in Parkin^Δ1–79^ levels. We hypothesize differences in cis- and trans-regulatory elements, ribosomal efficiency at the iTIS, an unidentified second hit, differences in polygenic scores, or a difficult-to-detect third *PRKN* variant to potentially impact Parkin^Δ1–79^ levels in patients with particularly young disease onset. Additionally, other unreported covariates such as environmental factors and comorbidities may also modify AAO in this specific group of PARK-*PRKN* patients.^43^ Interestingly, a single *PRKN*^delEx2^ variant and other pre-iTIS variants were not associated with a later AAO. Especially in case of compound heterozygous *PRKN*^delEx2^ carriers, we first hypothesized an effect of approximately half the observed shift seen in homozygous carriers. However, the smaller number of analyzed cases and potential effect of undocumented confounding factors may have limited the detection of such an effect. Besides that, our findings may indicate that gene dosage of Parkin^Δ1–79^-competent alleles such as *PRKN*^delEx2^ is crucial to affect AAO. Given our data, the unique feature of the *PRKN*^delEx2^ variant decreasing the distance between the regulatory 5’-untranslated region and the iTIS likely facilitates an increase of Parkin^Δ1–79^ levels. For other pre-iTIS variants, translation initiation may primarily occur at the canonical start codon, resulting in misfolded and unstable protein.^44^ Therefore, lower levels of internal ATG initiation likely result in reduced Parkin^Δ1–79^ quantity compared to *PRKN*^delEx2^ carriers. Thus, our finding complements previous research on genotype-phenotype correlation with this distinct case of Parkin^Δ1–79^ compensation in biallelic *PRKN*^delEx2^ carriers.^7^

While the presence of Parkin^Δ1–79^ (in addition to the more prominent, full-length Parkin isoform) in human cells and partial function of similar ΔUbl variants using overexpression models has been observed previously,^16–20, 22, 36, 45^ its endogenous activity, stability, and regulation in PD patient-derived material remained unclear. Western blot quantification experiments revealed increased levels of truncated Parkin^Δ1–79^, particularly in cells carrying a biallelic *PRKN*^delEx2^ variant but not in cells overexpressing *PRKN*^delEx2^ construct. In overexpression models, Parkin expression is driven by a strong promoter which likely masks effects observed in endogenous cell models, also reflected by a lack of measurable Parkin autoubiquitination and degradation with prolonged mitochondrial depolarization. Furthermore, Parkin^Δ1–79^ levels were not significantly increased in a homozygous *PRKN*^c.100_101insC^ cell model. These patterns likely indicate the importance of closer proximity of the iTIS to the transcripts 5’ end in *PRKN* Exon 2 deletion carriers to further enhance Parkin^Δ1–79^ translation efficiency. *In silico* TIS prediction further supports our *in vitro* protein quantification results. Here, the high probability score predicted for the internal TIS matches the increased Parkin^Δ1–79^ levels measured in *PRKN*^delEx2^ cell models. However, despite 9%-55% of total Parkin expressed in these cells being Parkin^Δ1–79^, the internal TIS in both wild-type and c.100_101insC *PRKN* was predicted at probability scores below the default threshold (0.01) by TIS Transformer. Thus, although evidently capable of correctly predicting multiple TIS per analyzed transcript, a lack of annotated multi-TIS transcripts in the Ensemble data used for training the transformer network may indeed limit model performance in this particular case.^42^ Nonetheless, high probability outputs not only for the *PRKN* Exon 2 deletion transcript but also start codon missense variants such as c.2T>C, emphasize the additional utility of optimized deep learning models for auxiliary analysis of start-loss or frameshift variants and their effect on remaining expression of N-terminally truncated proteins via alternative internal TIS such as in Parkin^Δ1–79^.

Furthermore, unlike full-length Parkin levels, Parkin^Δ1–79^ quantities remained relatively stable with prolonged mitochondrial membrane depolarization. Full-length Parkin self-regulates by autoubiquitination and subsequent proteasome degradation.^46^ A decreased ubiquitin ligase activity and mitochondrial recruitment of Parkin^Δ1–79^ may impair autoubiquitination. Furthermore, previous affinity purification-mass spectrometry experiments suggested a successful association of a ΔUbl-Parkin fragment similar to Parkin^Δ1–79^ with the outer mitochondrial membrane but not the proteasome.^47^ Thus, lack of the Ubl-domain may impair proteasomal association and degradation of Parkin, stabilizing it.

Previous functional assessments of Parkin^Δ1–79^ and closely related ΔUbl-Parkin fragments focused on assays relying on overexpression experiments.^16, 17^ Complementing these findings, we provide multi-level evidence for endogenous Parkin^Δ1–79^ function in patient-derived neuronal models of an asymptomatic biallelic *PRKN*^delEx2^ carrier^21^ and CRISPR-Cas9-edited isogenic cell lines, as well as neuroblastoma cells harboring a pre-internal TIS frameshift variant. Endogenous Parkin^Δ1–79^ is capable of substrate ubiquitination, PINK1-dependent mitochondrial membrane translocation, and mitophagy initiation. This is in agreement with previous findings showing impaired but still remaining mitochondrial recruitment and activity of ΔUbl-Parkin.^10, 16, 38^ The residual activity occurs despite lacking the Ubl domain and, thus, its phosphorylation site, which facilitates efficient mitochondrial recruitment and Parkin activation via PINK1 in full-length Parkin.^48^ Previous studies demonstrated that phospho-ubiquitin alone is sufficient for the recruitment and activation of Parkin.^17^ Thus, phospho-ubiquitin accumulation on the mitochondrial membrane substitutes for the phospho-Ubl domain to displace the catalytic RING2 domain from RING0 and activates Parkin^Δ1–79^. Of note, the efficiency of substrate ubiquitination by Parkin^Δ1–79^ was much more comparable to full-length Parkin in our overexpression models than in cell models endogenously expressing Parkin. Here, the manifold abundance of Parkin by overexpression may alleviate the reduced recruitment efficiency of Parkin^Δ1–79^ to the outer mitochondrial membrane. Importantly, the presence and activity of Parkin^Δ1–79^ in patients with pre-internal TIS *PRKN* variants adds further complexity to studies of specific genetic variants and the generation of gene knockouts. Residual amounts of Parkin^Δ1–79^ may confound results, and care should also be taken to inactivate its expression for induction of a complete gene knockout. Furthermore, besides *PRKN*, numerous human genes have been shown to transcribe multiple protein isoforms initiated at alternative start codons within a common reading frame.^49–52^ Here, the underlying molecular mechanism and disease relevancy can be quite different compared to Parkin^Δ1–79^. In contrast to Parkin^Δ1–79^, these alternative proteoforms are transcribed from TIS upstream of the canonical start codon rather than downstream. Interestingly, in the case of the Spastic Paraplegia 4 causing gene *SPAST*, a gain-of-toxicity disease mechanism linked to the less abundant M1-spastin initiated upstream of the canonical M87-spastin has been hypothesized.^52^

Recent progress in the development and characterization of THPP small molecule allosteric modulators of Parkin has demonstrated the capability of these compounds to boost a Ubl-independent Parkin activation mechanism selectively and constitutes a promising future therapeutic opportunity.^22, 23^ We here provide first evidence for BIO-2007817 boosting endogenous Parkin’s^Δ1–79^ ubiquitin ligase activity and turnover in patient-derived dopaminergic midbrain neurons. Importantly, THPP treatment decreased ubiquitination activity in healthy controls. Whereas this opposing effect has so far not been observed in exogenous expression models,^22, 23^ we hypothesize that by boosting the less efficient pUb-dependent activation of Parkin over pUbl-dependent activation, full-length Parkin translocation, and ubiquitination efficiency may become impaired upon high dosage THPP treatment under fully endogenous conditions. This observation of opposing, variant-selective modulation by BIO-2007817 warrants further replication to avoid potential adverse effects in individuals expressing Parkin with an intact Ubl-domain. Most importantly, our data further support the potential of THPP compounds in future personalized treatment strategies for patients with residual Parkin^Δ1–79^ expression or defective Parkin Ubl domains. They may further benefit from deubiquitination inhibitors such as MTX325, which, in contrast to THPPs, is already being examined in a phase 1 clinical trial.^53^

In summary, our data provide translational evidence for residual Parkin function in a subset of PARK*-PRKN* patients. These findings provide additional functional insight into the Parkin^Δ1–79^ protein fragment and constitute a potential avenue for future pharmacological intervention in these patients. In this regard, small molecule positive allosteric modulators of Parkin may particularly benefit patients with residual Parkin^Δ1–79^ expression, such as biallelic *PRKN*^delEx2^ carriers. Our study warrants further in-depth research on residual Parkin^Δ1–79^ in variant carriers other than biallelic *PRKN*^delEx2^ carriers, i.e., to examine the potential of boosting expression and response to THPP compounds as well as deubiquitination inhibitors. In addition, the impact of Parkin^Δ1–79^ level differences and their potential upstream causes on the AAO range observed in biallelic *PRKN*^delEx2^ carriers remains to be elucidated. Importantly, besides *PRKN*, other disease-linked genes may produce truncated but partially functional proteins by using internally initiated translation with an effect on the disease phenotype.

## Data Availability

All data presented and analyzed in this study are available from the corresponding authors upon reasonable request.

## Acknowledgments

We thank all patients and their families for participating in this study. C.M.S. is a NHMRC Practitioner Fellow (APP1103688).

## Data availability

To prevent identification, individual age at onset and age at examination data are reported as ranges, not exact ages. Readers who wish to reproduce the findings presented in this study will find the precise information of previously reported individuals in the referenced literature and the MDSGene database or may contact the corresponding authors to request full access.

