## Supplemental Information for "Usage of an Alternative Translation Start Site in *PRKN* mutation carriers: Key to Later-Onset Parkinson’s Disease and a Novel Therapeutic Target"

### Supplementary Methods

#### Cell culture

SH-SY5Y neuroblastoma cells, human embryonic kidney cells (HEK 293FT, for reprogramming-related virus production), and primary human dermal fibroblasts were cultivated in Dulbecco's Modified Eagle Medium (Gibco) containing 10% fetal bovine serum and 1% penicillin/streptomycin using standard procedures. Cells were passaged at 80%-90% confluence. Passage numbers of harvested human dermal fibroblasts did not exceed 15 and did not vary by more than two.

To disrupt the mitochondrial membrane potential, SH-SY5Y cells, human dermal fibroblasts, and dopaminergic midbrain neurons were treated with 1 µM valinomycin (Sigma-Aldrich) for 3, 6, or 14 hours, respectively.

Further, hiPSCs were cultured on Matrigel-coated plates and maintained in mTeSR Plus cell culture medium. hiPSCs were passaged manually at 70-80% confluence.

#### Viral vectors and transduction

Lentiviral expression plasmids for Parkin-wt, Parkin-delEx2, and Parkin-c.1-237del were generated by cloning cassettes containing the respective cDNA sequences, followed by an internal ribosomal entry site (IRES), and the puromycin N-acetyl-transferase (PAC) gene into NK57 lentiviral vectors.

Viral particles were produced in HEK 293FT cells by transfection with target, VSV-G, and 8.2ΔR plasmids using FuGENE HD (Promega) transfection reagent. Isogenic SH-SY5Y-wt and SH-SY5Y-PRKN-KO neuroblastoma cells were transduced with each lentiviral construct for 48h.

#### Induced pluripotent stem cell generation and characterization

The analyzed healthy control cell lines SFC156-03-01 (STBCi101-A) and SFC163-03-01 (STBCi102-A), which also served as parental lines for the isogenic *PRKN*$^{delEx2}$ and *PRKN*$^{delEx3}$ lines, have been reported previously.[1,2] Fibroblast cultures from patient B-300 and the unaffected *PRKN*$^{delEx2}$ carrier L-4985 were transduced to overexpress cMYC, KLF4, and KOS using the CytoTune-iPS 2.0 Sendai Reprogramming Kit (Thermo Fisher Scientific) according to the manufacturer's instructions. The newly generated hiPSC and isogenic *PRKN*$^{delEx2}$ and *PRKN*$^{delEx3}$ cell lines were characterized to ensure correct genotype, pluripotency, clearance of Sendai virus components, and absence of mycoplasma contamination as previously described (Supplementary Fig. 2).[3,4] In brief, embryoid bodies (EBs) were differentiated from each iPSC line by transferring cells to Ultra-Low Attachment 6-well plates (Corning) containing TeSR-E8-Medium containing 4mg/ml PVA (Sigma-Aldrich). After 2 days, the culture medium was changed to E6-Medium (gibco). EBs were maintained in E6 for 14 days, changing the culture medium every other day. RNA was isolated from fresh iPSC and EB pellets (500 x g, 5 min) using the RNeasy Mini Kit (Qiagen). The RNA was reversed transcribed with the First Strand cDNA Synthesis Kit (Thermo Fisher Scientific). The absence of Sendai virus components and mycoplasma contamination were confirmed by PCR. Here, fibroblasts 5 days after transduction and a mycoplasma-positive sample served as positive controls. To assess the expression of pluripotency markers *GDF3*, *NANOG*, *OCT4*, *SOX2* in hiPSCs and germ layer markers *GATA4*, *MSX1*, *MYH6*, *NCAM*, *PAX6*, *RUNX1*, and *SOX17* in EBs, quantitative real-time PCR runs were conducted with Maxima SYBR Green (Thermo Fisher Scientific) on the Lightcycler 96 (Roche). Additionally, iPSCs were fixed using 4% PFA and immunohistochemically stained for the pluripotency markers OCT4, NANOG, SSEA-4, and TRA-1-60. Furthermore, *PRKN* cDNA transcribed from iDNs was amplified from Exon 1(Primer F: GCGCGGCTGGCGCCGCTGGCGGCA) to Exon 10 (Primer R: GCTTCTTTACATTCCCGGCAG), run on an agarose gel and analyzed by Sanger sequencing to confirm the *PRKN* genotype for each cell line. Additional primers to determine *PRKN* expression levels by quantitative real-time PCR were Exon 9F (TGACCAGAGGAAAGTCACCTG) and Exon 11R (CAGGGCTTGGTGGTTTTCTTG). All primers and antibodies not explicitly mentioned have been reported previously.[3,4]

#### Genome-edited cell lines

We used several genome-edited neuroblastoma and hiPSC lines, including a previously generated isogenic SH-SY5Y cell line lacking PINK1 (SH-SY5Y *PINK1*$^{KO}$).[5] Further, SH-SY5Y *PRKN*$^{c.100\_101insC}$ cells, SH-SY5Y *PRKN*$^{KO}$ (deletion of Exon 3), isogenic iPSC *PRKN*$^{delEx2}$, and iPSC *PRKN*$^{delEx3}$ cells were newly generated using an RNA-guided CRISPR/Cas9 endonuclease following previously established protocols.[6,7] For the SH-SY5Y *PRKN*$^{c.100\_101insC}$ line, cells were transiently transfected with episomal vectors expressing both a human codon-optimized Cas9 and a guide RNA (gRNA) containing a 20-base long sequence that matches the human *PRKN* target sequence 5'- GTGGTTGCTAAGCGACAGG-3' in Exon 2. For iPSC *PRKN*$^{delEx2}$ we used two gRNAs targeting 5'-TGTCAGGTTCAACTCCAGCC-3' (in Exon 2) and 5'-GCAGGTGAGTCTCCCTTGG-3' on the junction of Exon 2 and Intron 2-

3. To generate SH-SY5Y *PRKN*$^{KO}$ and iPSC *PRKN*$^{delEx3}$, we used two gRNAs targeting 5'-TCAGCAGCTCAGTCCTCCC-3' (in Exon 3) and 5'- AGCTGGAAGTCCAGGTAAT-3' on the junction of Exon 3 and Intron 3-4. Upon transfection, cells were resuspended in the corresponding growth medium, counted, and plated onto Petri dishes at a density of 1 cell/cm2. Cells were grown until they formed distinct, monoclonal colonies. The colonies were scraped off, transferred into different wells of a 6-well plate, and propagated to obtain enough material for DNA extraction. All variants were confirmed by Sanger sequencing.

#### Mitochondrial isolation

Fractionation of SH-SY5Y cells was conducted as previously described.[8,9] Immediately after collection, cells were homogenized in an isolation medium containing 250 mM sucrose, 10 mM Tris-HCl, and 1 mM EDTA at pH 7.4. Homogenates were centrifuged for 20 min at 1,500 x g. Supernatants were transferred to a fresh tube and centrifuged for an additional 10 min at 12,000 x g. Supernatants containing cytosolic fractions were transferred to a fresh tube. Pellets containing the mitochondria were resuspended in Radioimmunoprecipitation assay (RIPA) buffer comprised of 25 mM Tris-HCl, 150 mM NaCl, 1% NP-40, 1% sodium deoxycholate, and 0.1% SDS at pH 7.6 supplemented with cOmplete protease and phosSTOP phosphatase inhibitors (Roche). Proteins in cytosolic fractions were concentrated with Amicon Ultracel 10K centrifugal filters (Millipore). Subsequently, fractions were analyzed by western blotting.

#### *In silico PRKN* translation initiation site prediction

cDNA sequences of wild-type (NM_004562.3), delEx2, c.100_101insC, and c.2T>C *PRKN* variants were analyzed using TIS Transformer (Version 1.0) in conjunction with the transcript-transformer python package (Version 0.4).[10] Minimum prediction threshold was set to 0.001 to track model prediction outputs for the internal TIS across all analyzed sequences.

### Supplementary Tables

**Supplementary Table 1 Demographics of variant carriers reported on MDSGene and included in the AAO analysis propensity score matched to respective homozygous *PRKN*$^{delEx2}$ variant carriers in Table 1**

| ID | Center | Country | Sex | AAO Range | AAE Range | Previously Reported | 1$^{st}$ Variant | 2$^{nd}$ Variant | Match Subclass |
|---|---|---|---|---|---|---|---|---|---|
| 1i | Nara Medical U | Japan | Male | 31-35 | 41-45 | Yes[11] | delEx2 | delEx4 | 8 |
| 1i | Juntendo U | Japan | Male | 46-50 | 56-60 | Yes[12] | delEx3 | delEx3 | 15 |
| II-2 | Juntendo U | Japan | Male | 21-25 | 41-45 | Yes[13] | delEx3 | delEx3 | 12 |
| I-1 | Juntendo U | Japan | Male | 16-20 | NA | Yes[13] | delEx3 | delEx3 | 11 |
| patient 2 | Tokushima U | Japan | Female | 16-20 | 56-60 | Yes[14] | delEx3-4 | delEx3-4 | 14 |
| M5 | Zhejiang U | China | Female | 16-20 | 26-30 | Yes[15] | delEx3-4 | delEx3-4 | 20 |
| IV-4 | Niigata U | Japan | Female | 16-20 | 31-35 | Yes[16] | delEx4 | delEx4 | 16 |
| II-1 | Juntendo U | Japan | Male | 16-20 | NA | Yes[17] | delEx4 | delEx4 | 10 |
| 479-10 | Mersin U | Turkey | Male | 16-20 | 31-35 | Yes[18] | delEx4 | delEx4 | 26 |
| IV-3 | Niigata U | Japan | Male | 11-16 | 36-40 | Yes[16] | delEx4 | delEx4 | 6 |
| IV-6 | Niigata U | Japan | Female | 6-10 | 26-30 | Yes[16] | delEx4 | delEx4 | 9 |
| II-1 | Niigata U | Japan | Male | 26-30 | 31-35 | Yes[16] | delEx6-7 | delEx6-7 | 5 |
| 13 | Juntendo U | Japan | Male | 41-45 | 56-60 | Yes[19] | delEx2-4 | delEx2-4 | 25 |
| III-1 | Istanbul U | Turkey | Male | 36-40 | 51-55 | Yes[20] | c.1084-1delG | c.1084-1delG | 17 |
| C2 | Juntendo U | China | Female | 16-20 | 31-35 | Yes[21] | c.1321T>C | p.Ala138Glyfs*7 | 23 |
| III:1 | Cantabria U | Spain | Male | 26-30 | NA | Yes[22] | c.1334G>A | c.1334G>A | 4 |
| PK-21 | Barcelona U | Spain | Male | 41-45 | 51-55 | Yes[23] | c.155delA | c.155delA | 1 |
| 8 | IDIBAPS | Spain | Male | 36-40 | 56-60 | Yes[24] | c.155delA | c.155delA | 2 |
| II-2 | HPPH | China | Female | 21-25 | 56-60 | Yes[25] | c.619-1G>C | delEx1-2 | 19 |
| II-7 | HPPH | China | Male | 21-25 | NA | Yes[25] | c.619-1G>C | delEx1-2 | 22 |
| II:1 | Central South U | China | Female | 21-25 | 26-30 | Yes[26] | c.850G>C | c.850G>C | 24 |
| II:3 | Central South U | China | Male | 6-10 | 21-25 | Yes[26] | c.850G>C | c.850G>C | 18 |
| II:1 | Shandong U | China | Male | 26-30 | 36-40 | Yes[27] | c.850G>C | delEx6 | 21 |
| M6 | Zhejiang U | China | Male | 21-25 | 36-40 | Yes[15] | delEx2 | delEx6 | 3 |
| A2 | Juntendo U | Japan | Male | 11-15 | 51-55 | Yes[21] | p.Thr175Profs*2 | p.Thr175Profs*2 | 7 |
| A3 | Juntendo U | Japan | Female | 11-15 | 36-40 | Yes[21] | p.Thr175Profs*2 | p.Thr175Profs*2 | 13 |

AAE = age at examination; U = University; IDIBAPS = Institut d'Investigacions Biomèdiques August Pi i Sunyer; HPPH = Henan Provincial People's Hospital.

**Supplementary Table 2 Human dermal fibroblasts analyzed in Supplementary Fig. 7**

| ID | AAE Range | Sex | Affected | *PRKN* Variant | Zygosity | Pathogenicity |
|---|---|---|---|---|---|---|
| L4985 | 66-70 | Female | No | delEx2 | Hom | Definitely pathogenic |
| B300 | 41-45 | Female | Yes | delEx7 | Hom | Definitely pathogenic |
| L3048 | 56-60 | Male | Yes | delEx4; c.823C>T | Compound het | Definitely pathogenic |
| L5415 | 31-35 | Female | Yes | c.823C>T; c.1054T>C | Compound het | Definitely pathogenic |
| L3244 | 41-45 | Female | Yes | del Ex1; c.823C>T | Compound het | Definitely pathogenic |
| L6069 | 71-75 | Male | No | WT | NA | NA |
| L6004 | 61-65 | Female | No | WT | NA | NA |

AAE = age at examination

**Supplementary Table 3 iPSC lines differentiated into midbrain dopaminergic neurons (iDN)**

| ID | AAE Range | Sex | Affected | *PRKN* variant | Zygosity | Pathogenicity | Isogenic |
|---|---|---|---|---|---|---|---|
| L4985-7 | 66-70 | Female | No | delEx2 | Hom | Definitely pathogenic | NA |
| B300-11 | 41-45 | Female | Yes | delEx7 | Hom | Definitely pathogenic | NA |
| SFC156-03-01 | 71-75 | Male | No | WT | NA | NA | Parental |
| SFC156-03-01delEx2 | 71-75 | Male | NA | delEx2 | Hom | Definitely pathogenic | Yes |
| SFC156-03-01delEx3 | 71-75 | Male | NA | delEx3 | Hom | Definitely pathogenic | Yes |
| SFC163-03-01 | 61-65 | Male | No | WT | NA | NA | Parental |
| SFC163-03-01delEx2 | 61-65 | Male | NA | delEx2 | Hom | Definitely pathogenic | Yes |

AAE = age at examination

**Supplementary Table 4 Demographics of patients with a heterozygous *PRKN*$^{delEx2}$ variant for *PRKN* (NM_004562.3) reported on MDSGene and included in the AAO analysis**

| ID | Country | Sex | AAO Range | AAE Range | Previously Reported | 1st Variant | 2nd Variant |
|---|---|---|---|---|---|---|---|
| SPD-322-010 | France | Male | 16-20 | 51-55 | Yes[28] | delEx2 | 202_203delAG |
| SPD-169-003 | France | Male | 26-30 | 41-45 | Yes[29] | delEx2 | delEx3 |
| SPD-166-010 | France | Male | 41-45 | 41-55 | Yes[29] | delEx2 | delEx3-4 |
| Patient 4 | China | Female | 26-30 | 31-35 | Yes[30] | delEx2 | 202_203delAG |
| M6 | China | Male | 21-25 | 36-40 | Yes[15] | delEx2 | delEx6 |
| M335 | China | Female | 26-30 | 41-45 | Yes[15] | delEx2 | delEx7-9 |
| II:3 | China | Male | 36-40 | 41-45 | Yes[31] | delEx2 | c.951G>C |
| II:2 | China | Male | 31-35 | 41-45 | Yes[31] | delEx2 | c.951G>C |
| II-a | Greece | Female | 16-20 | NA | Yes[32] | delEx2 | delEx5-7 |
| FB_P1 | Iran | Female | 6-10 | 21-25 | Yes[33] | delEx2 | delEx3 |
| B-151 | Italy | Male | 16-20 | 31-35 | Yes[34] | delEx2 | delEx5 |
| 37.12 | South Africa | Male | 41-45 | NA | Yes[35] | delEx2 | delEx9 |
| 3 | Italy | Male | 31-35 | 66-70 | Yes[36] | delEx2 | delEx2-3 |
| 2 | Italy | Male | 21-26 | 61-65 | Yes[36] | delEx2 | delEx2-3 |
| 1i | South Korea | Male | 6-10 | 16-20 | Yes[37] | delEx2 | delEx4 |
| 1i | Japan | Male | 31-35 | 41-45 | Yes[11] | delEx2 | delEx4 |
| 1i | Serbia | Female | 16-20 | NA | Yes[38] | delEx2 | delEx3 |
| 19 | France | Male | 11-15 | 46-50 | Yes[39] | delEx2 | delEx3 |
| 18 | France | Female | 36-40 | 46-50 | Yes[39] | delEx2 | delEx3 |
| 17i | South Korea | Male | 26-30 | 31-35 | Yes[40] | delEx2 | delEx4 |

AAE = age at examination
