## Supplemental Figures for "Usage of an Alternative Translation Start Site in *PRKN* mutation carriers: Key to Later-Onset Parkinson’s Disease and a Novel Therapeutic Target"

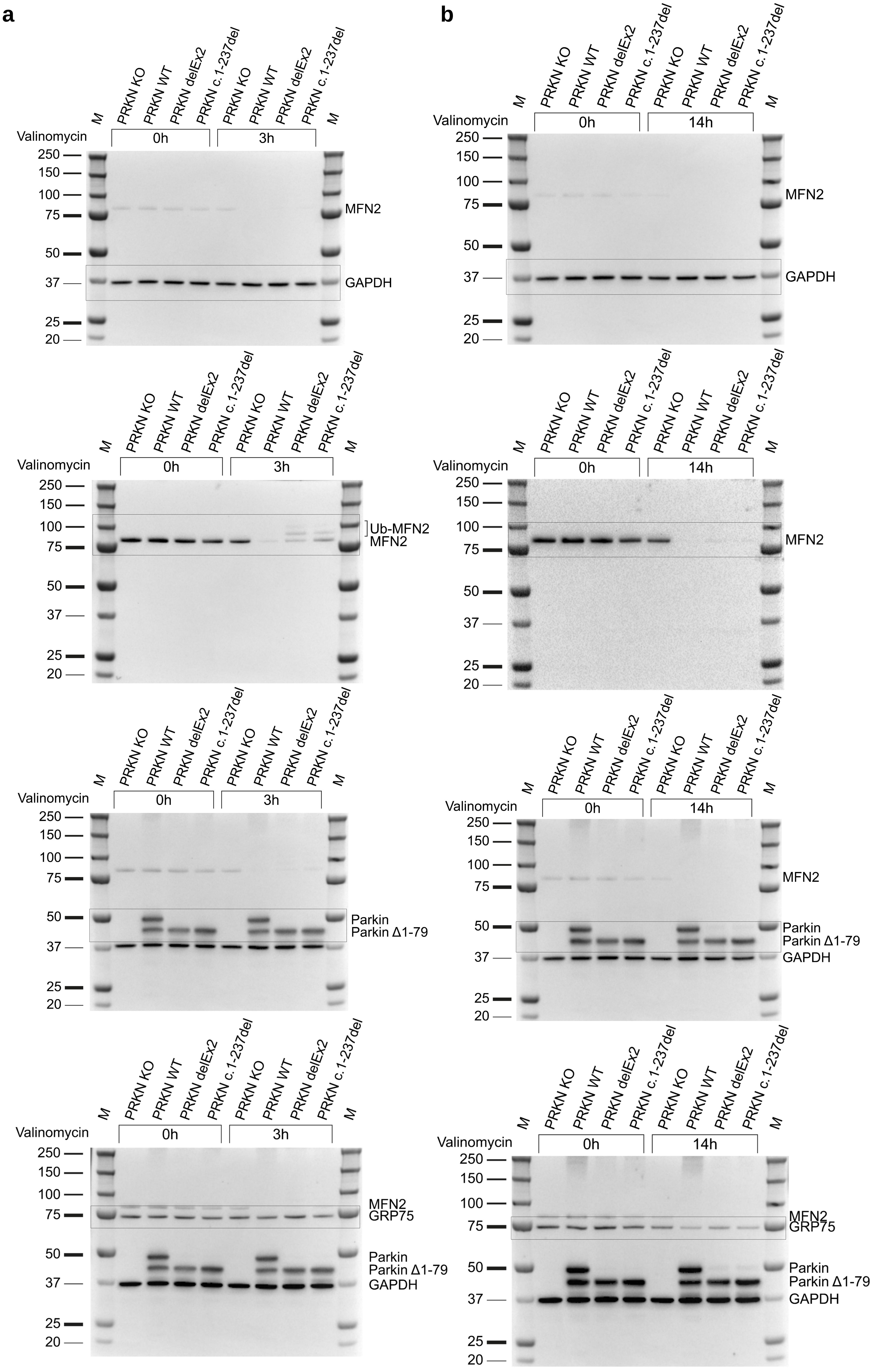


**Supplementary Figure 1: Complete western blots.** Full blots after 3 h (**a**) and 14 h (**b**) valinomycin treatment in SH-SY5Y cells overexpressing Parkin as shown in main figure 3. Blots are shown for each cropped signal, marked by boxes.

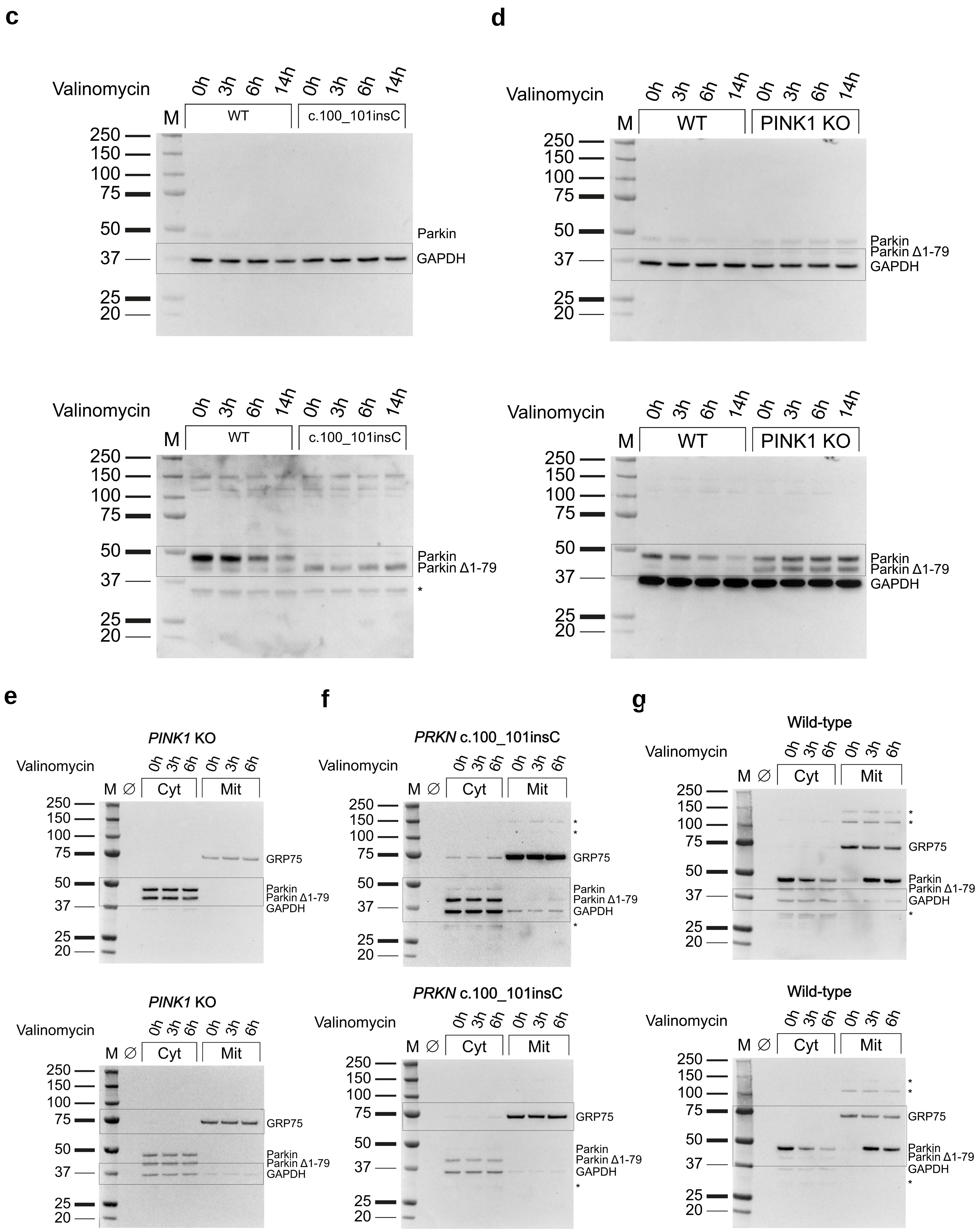


**Supplementary Figure 1: Complete western blots.** Full blots for c.100_101insC and wildtype (**c**) or PINK1 KO and wildtype cells (**d**) for quantification of endogenous Parkin in SH-SY5Y cells as well as full blots for the fractionation analysis of PINK1 KO (**e**) c.100_101inC (**f**) and wildtype (**g**) as shown in main figure 4. Blots are shown for each cropped signal, marked by boxes.

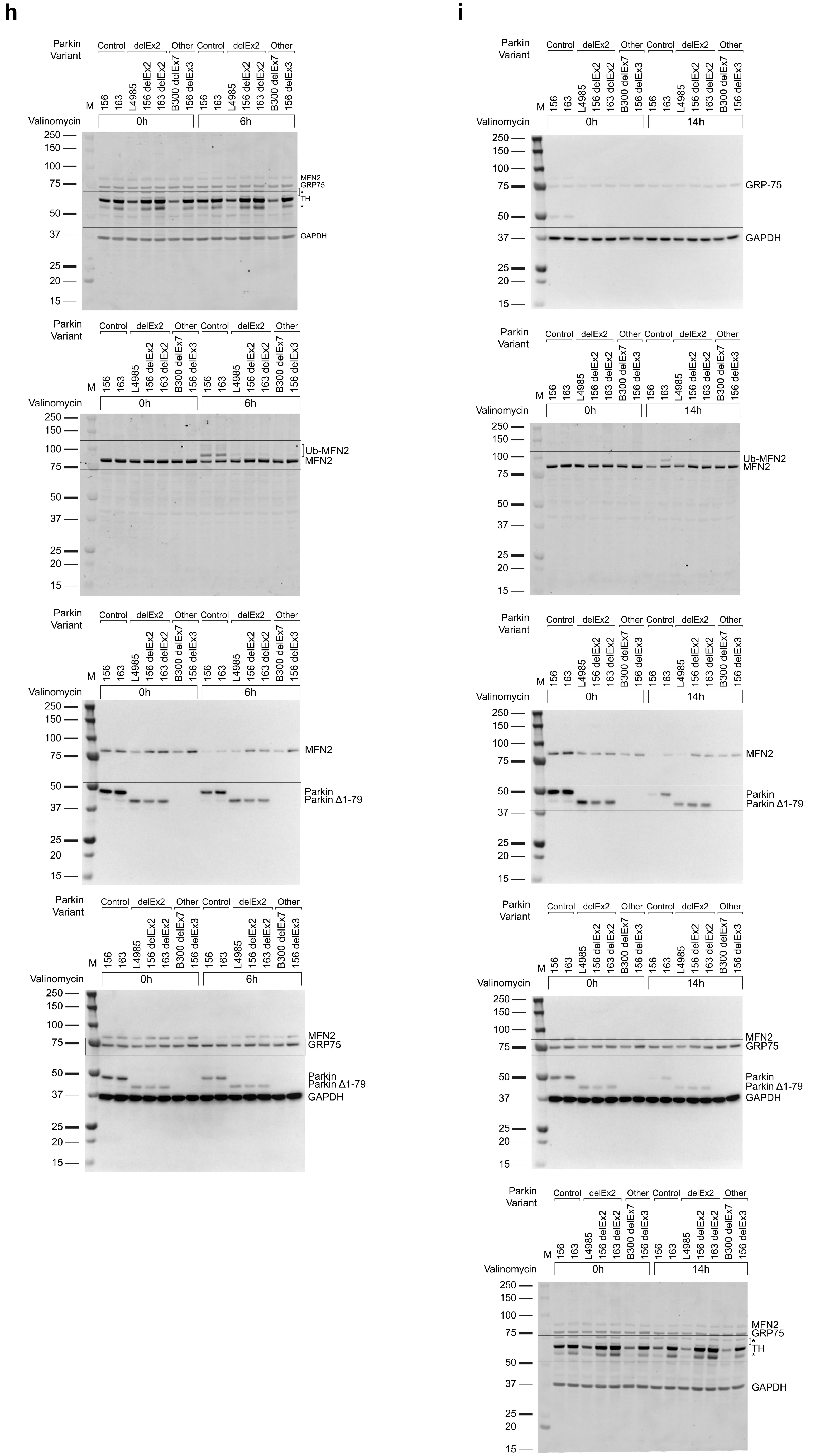


**Supplementary Figure 1: Complete western blots.** Full blots after 6 h (**h**) and 14 h (**i**) of Valinomycin treatment in iDNs as shown in main figure 5. Blots are shown for each cropped signal, marked by boxes.

**j**

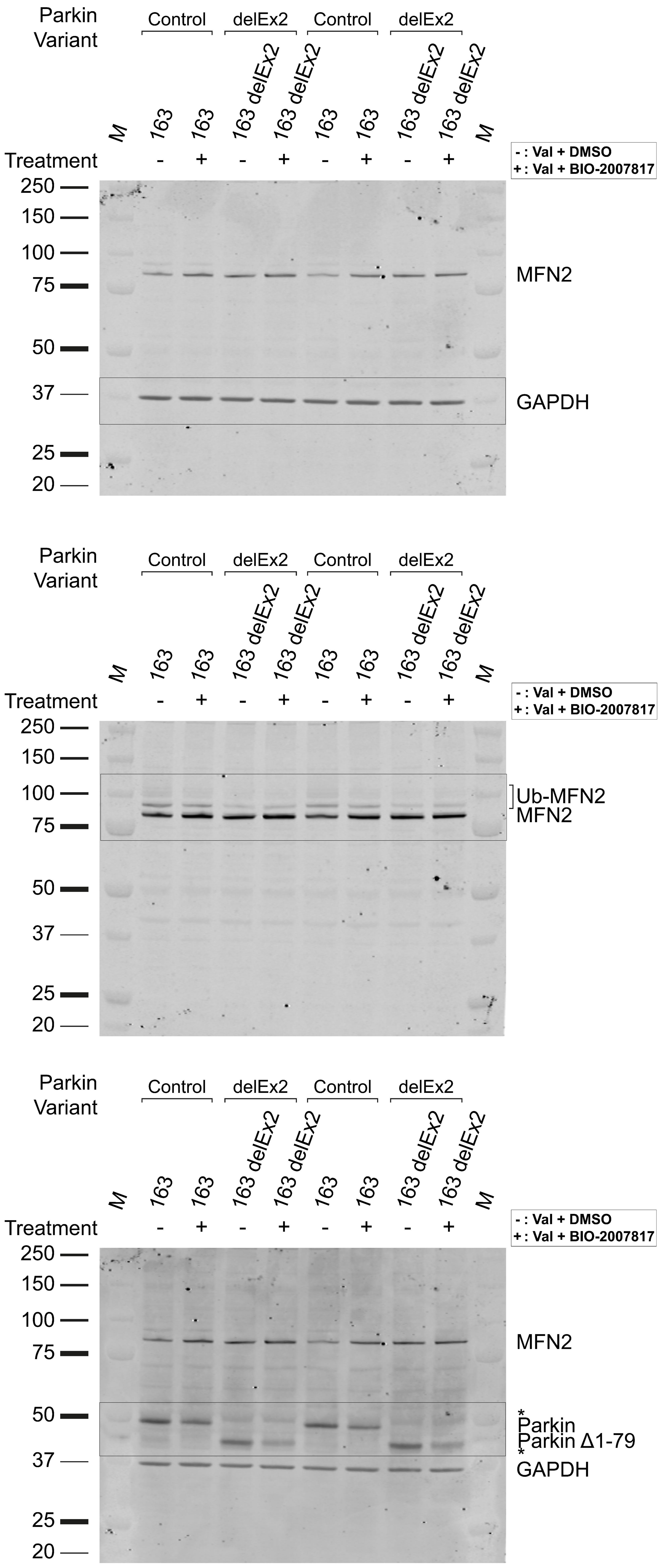


**k**

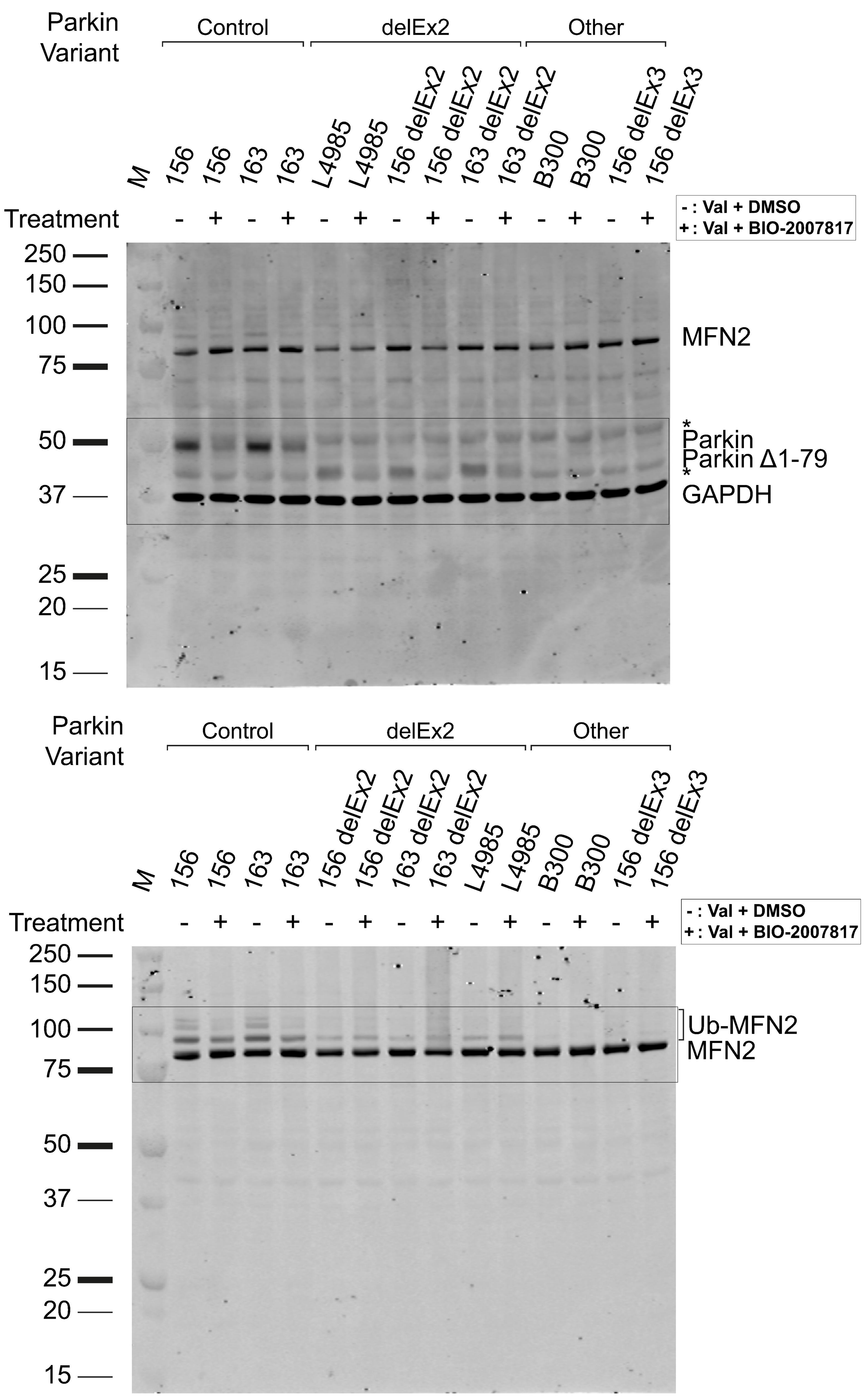


**Supplementary Figure 1: Complete western blots.** Full western blots of BIO-2007817 experiments in iDNs as seen in main figure 6A left (**j**) and right (**k**). Blots are shown for each cropped signal, marked by boxes.

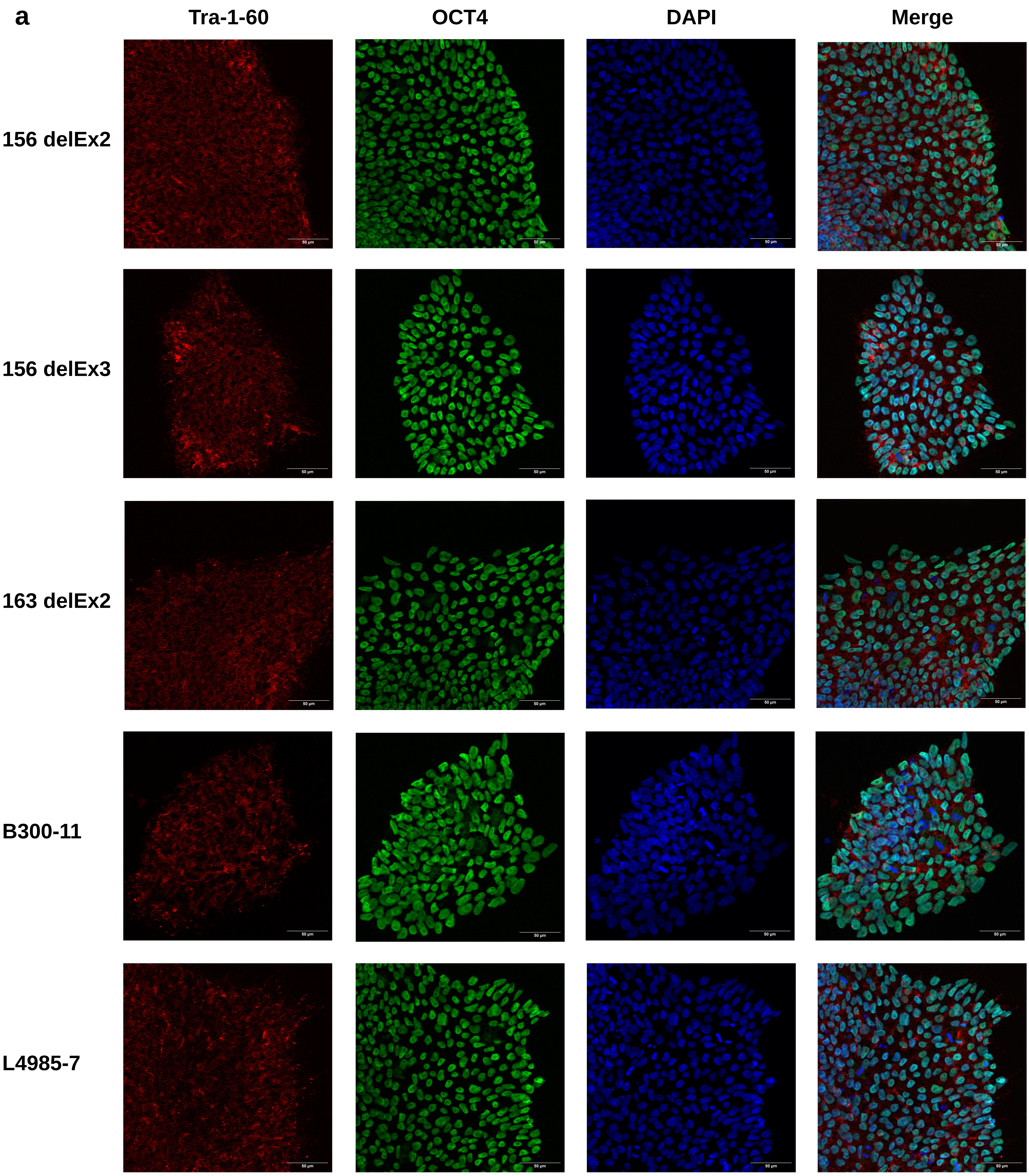


**Supplementary Figure 2a: hiPSC characterization.** Immunohistochemistry confocal microscopy images of all newly generated and CRISPR-Cas9 edited cell lines. Antibody staining for the pluripotency markers Tra-1-60 and OCT4. Nuclear counterstain via DAPI. Z-stacks were collapsed by average intensity projections.

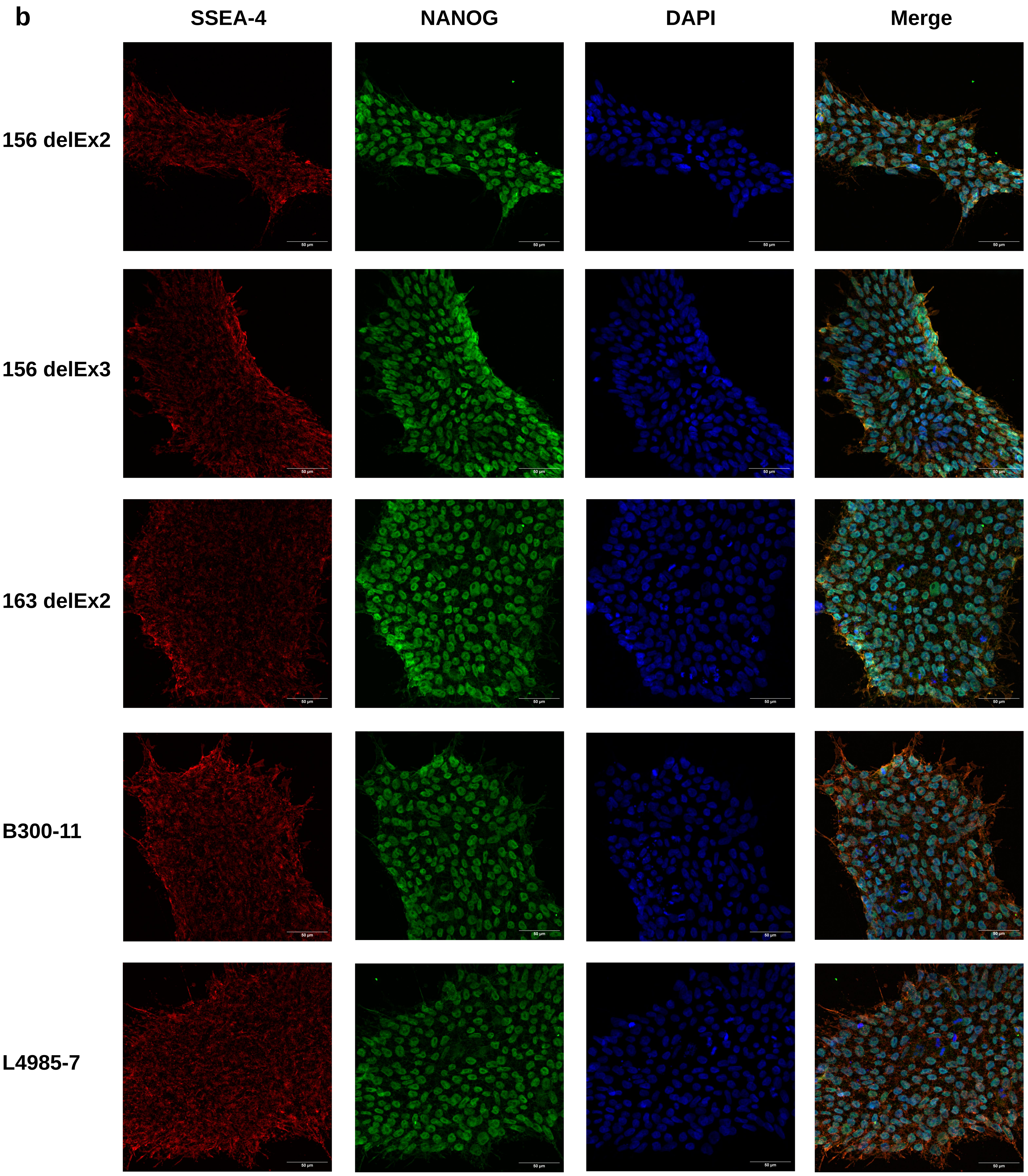


**Supplementary Figure 2b: hiPSC characterization.** Immunohistochemistry confocal microscopy images of all newly generated and CRISPR-Cas9 edited cell lines. Antibody staining for the pluripotency markers SSEA-4 and NANOG. Nuclear counterstain via DAPI. Z-stacks were collapsed by average intensity projections.

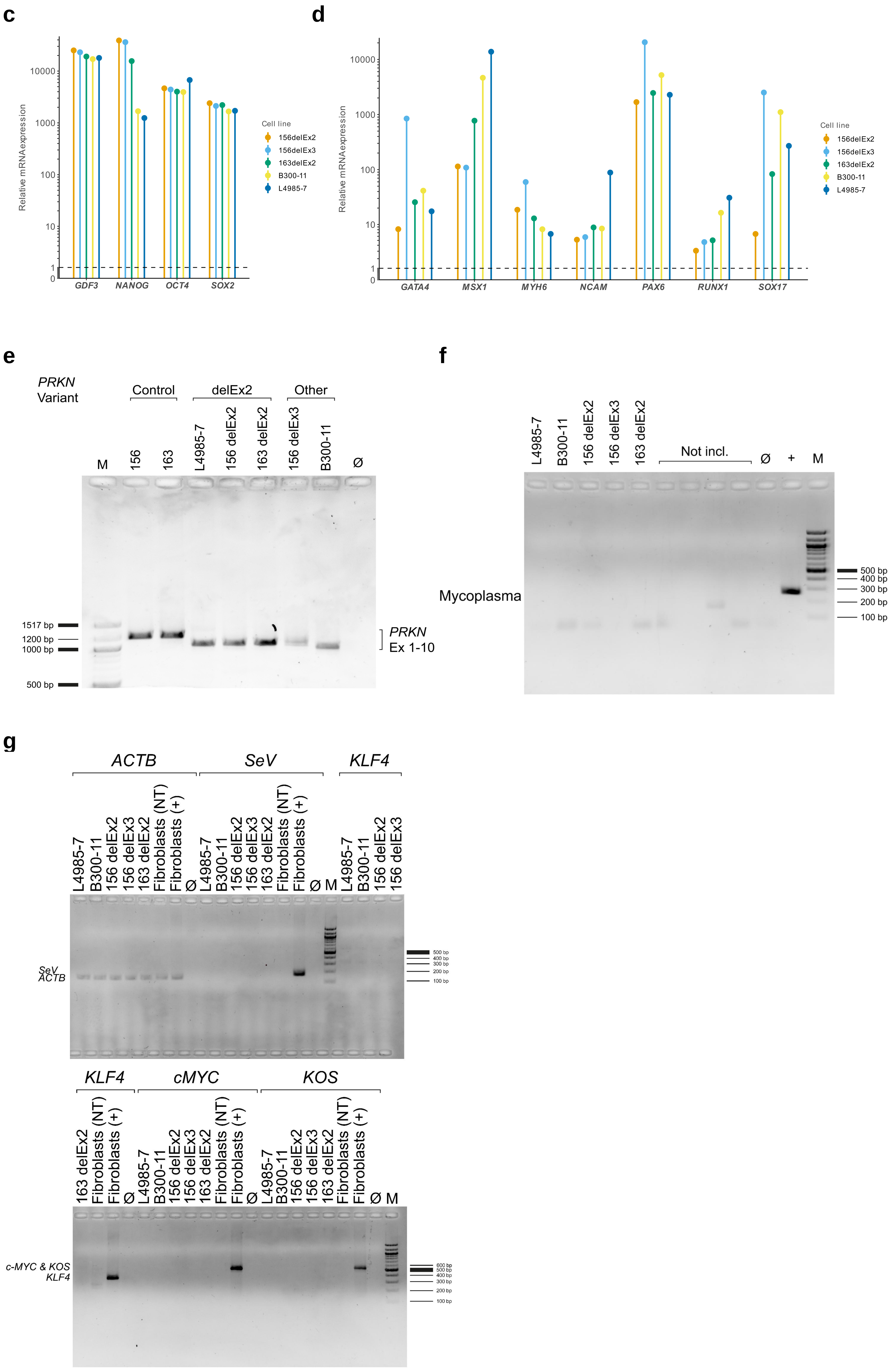


**Supplementary Figure 2: hiPSC characterization.** (**c**) Increased relative mRNA expression of the pluripotency markers *GDF3*. *NANOG*, *OCT*, and *SOX2* in all newly generated and CRISPR-Cas9 edited cell lines compared to Fibroblasts. (**d**) Increased relative mRNA expression of the ectodermal markers *NCAM* and *PAX6*, the mesodermal markers *MSX1*, *MYH6*, and *RUNX1* as well as the endodermal markers *GATA4* and *SOX17* in embryoid bodies differentiated from all newly generated and CRISPR-Cas9 edited cell lines compared to matched hiPSCs. (**e**) PCR amplification of *PRKN* Exon 1 to Exon 10 from iDN cDNA for all cell lines shown in main figure 5. (**f**) Mycoplasma PCR test of all newly generated and CRISPR-Cas9 edited cell lines. (**g**) PCR verifying the absence of Sendai virus reprogramming components for all newly generated and CRISPR-Cas9 edited cell lines.

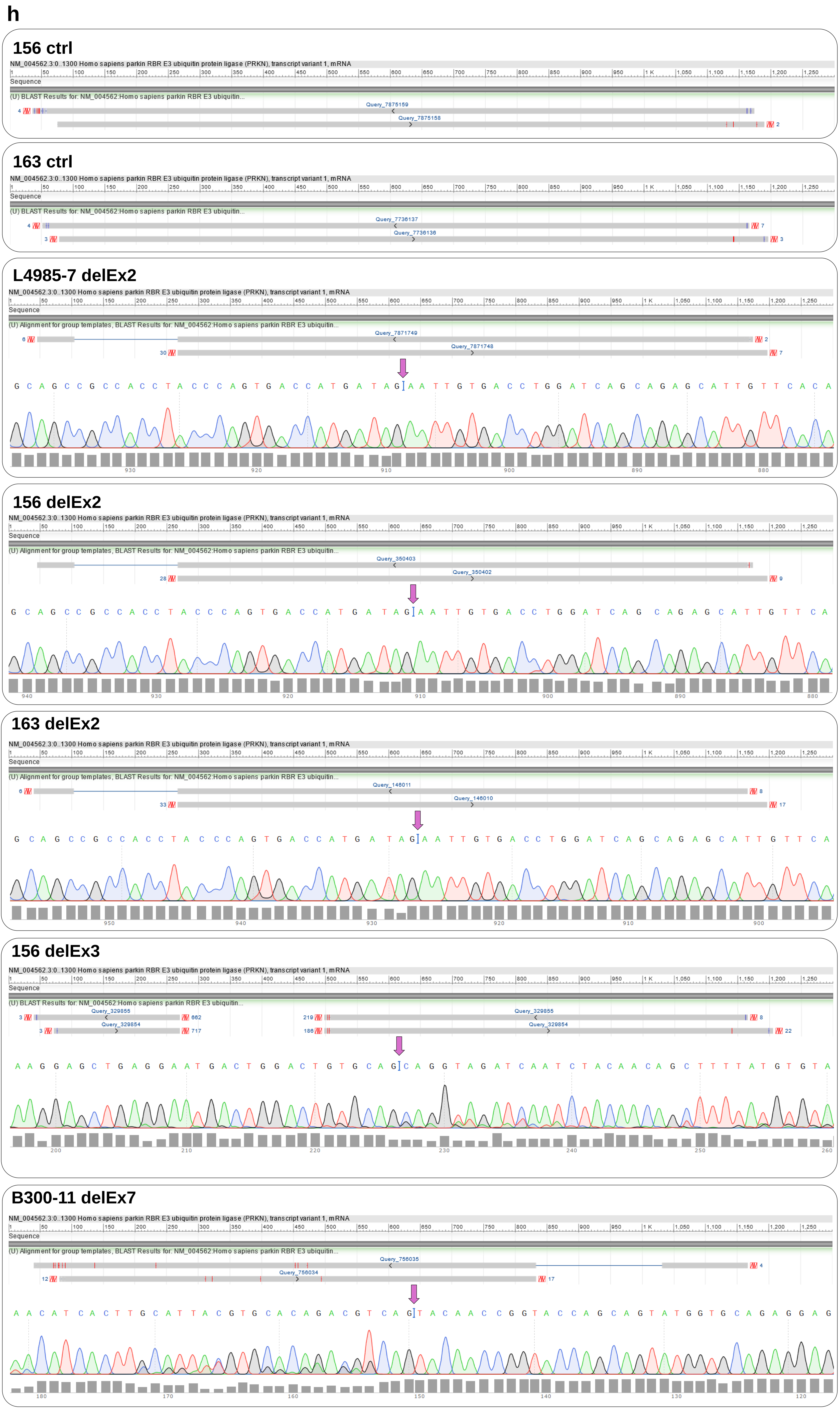


**Supplementary Figure 2h: hiPSC characterization.** Genotype confirmation by sanger sequencing using iDN cDNA amplified in (**e**). BLAST alignments to NM_004562.3 shown for all analyzed cell lines. Sequence windows of electropherograms relevant to each Exon deletion are shown. Arrows indicate the location of each variant. While patient B-300 is known to carry a homozygous deletion of Exon 7, disruption of the splicing context may result in the observed deletion of both Exon 7 & 8 on mRNA level.

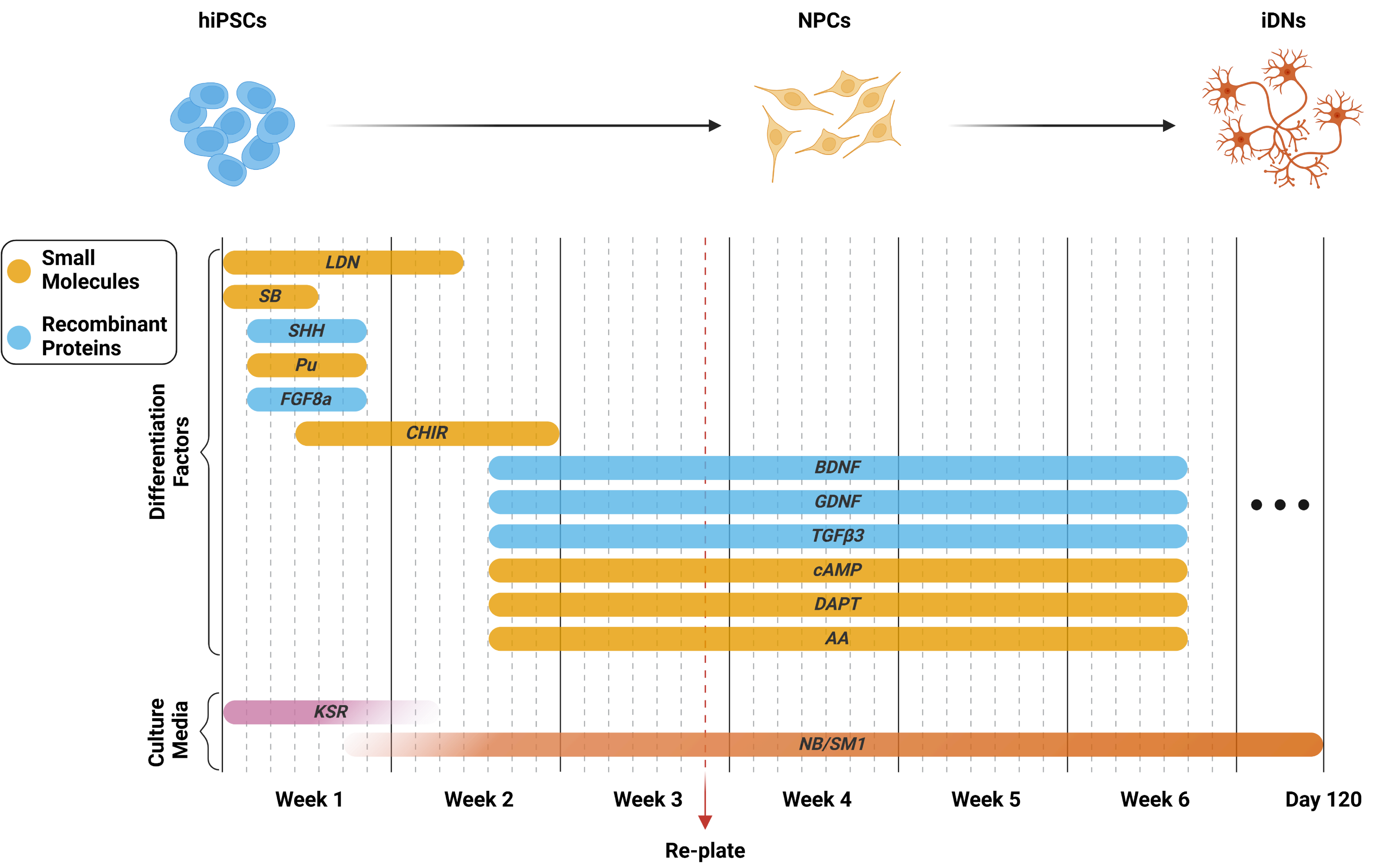


**Supplementary Figure 3: Differentiation scheme to generate midbrain dopaminergic neurons from patient-derived iPSCs.** Neuronal precursor cells (NPCs) were plated on PDL/LA coated cell culture plates on day 20, as indicated in red. iDNs were kept in culture until day 120. Created with BioRender.

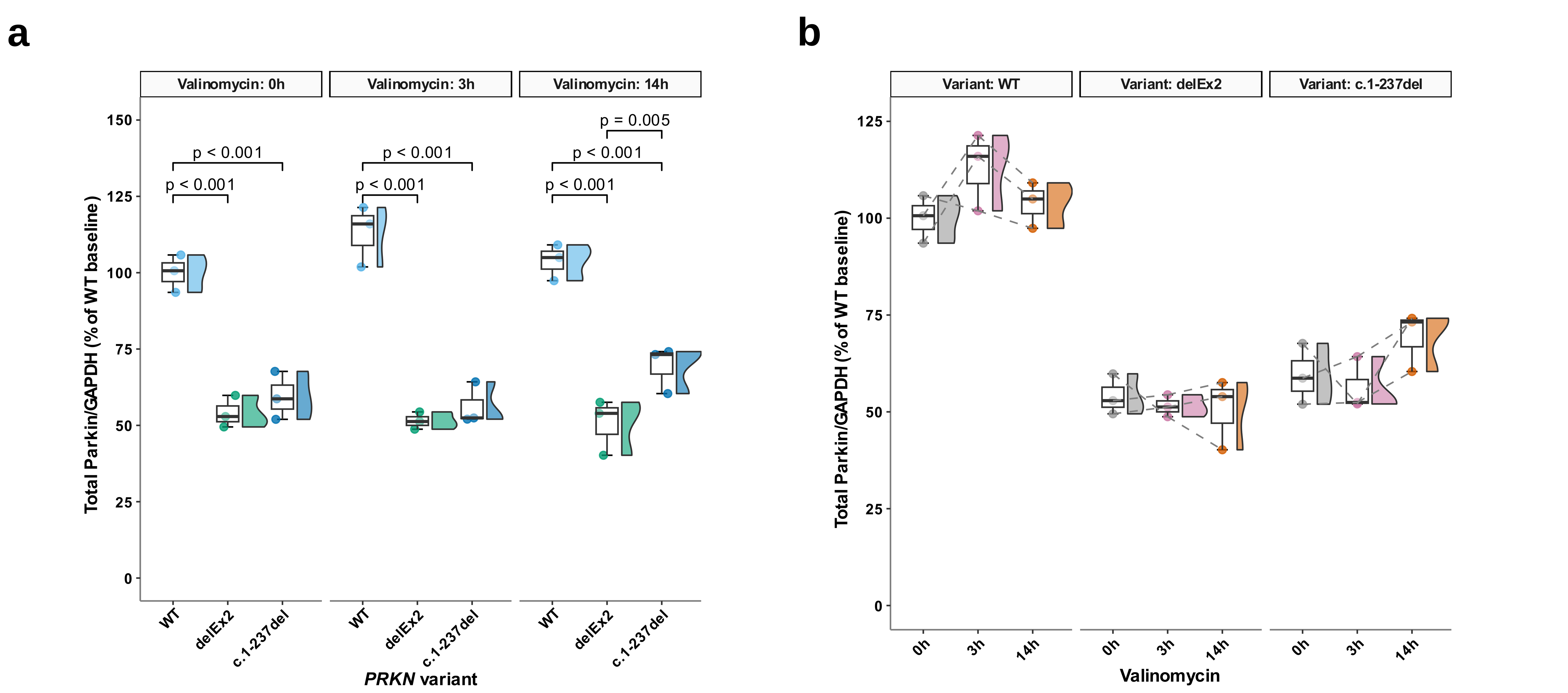


**Supplementary Figure 4:** Total Parkin analysis between groups (**a**) and within groups (**b**) complementary to overexpression experiments of three *PRKN* constructs in a *PRKN* knockout neuroblastoma cell model in SH-SY5Y cells shown in main figure 3. Sample size: n = 3 from independently repeated experiments across three cell passages. The significance threshold was set to p = 0.05. Whiskers extend to the largest and smallest values no further than 1.5 * IQR from the hinge. Pairwise comparisons of linear mixed effects model derived estimated marginal means were Holm-adjusted.

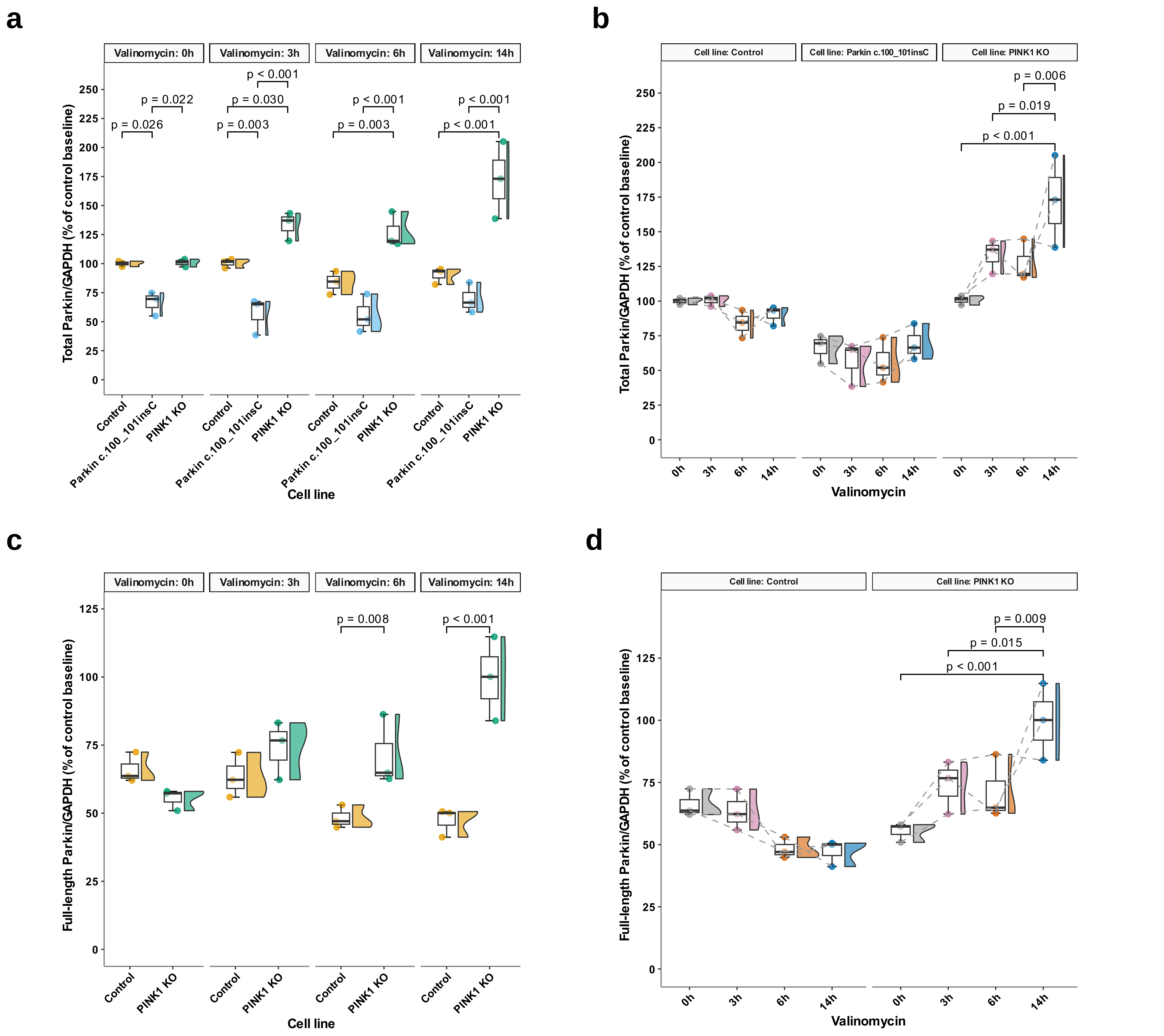


**Supplementary Figure 5:** (**a-b**) Endogenous full-length Parkin levels wild-type, *PRKN*$^{c.100\_101insC}$, and *PINK1*-KO SH-SY5Y neuroblastoma cells following mitochondrial depolarization with 1 μM valinomycin across multiple time points complementary to main figure 4. (**a-b**) Expression differences of total Parkin (full-length Parkin + Parkin$^{\Delta 1-79}$) between cell lines (**b**) and over increased durations of mitochondrial depolarization (**b**). Expression differences of only full-length Parkin between cell lines (**c**) and over increased durations of mitochondrial depolarization (**d**). Sample size: n = 3 from independently repeated experiments across three cell passages. The significance threshold was set to p = 0.05. Whiskers extend to the largest and smallest values no further than 1.5 * IQR from the hinge. Pairwise comparisons of linear mixed effects model derived estimated marginal means were Holm-adjusted.

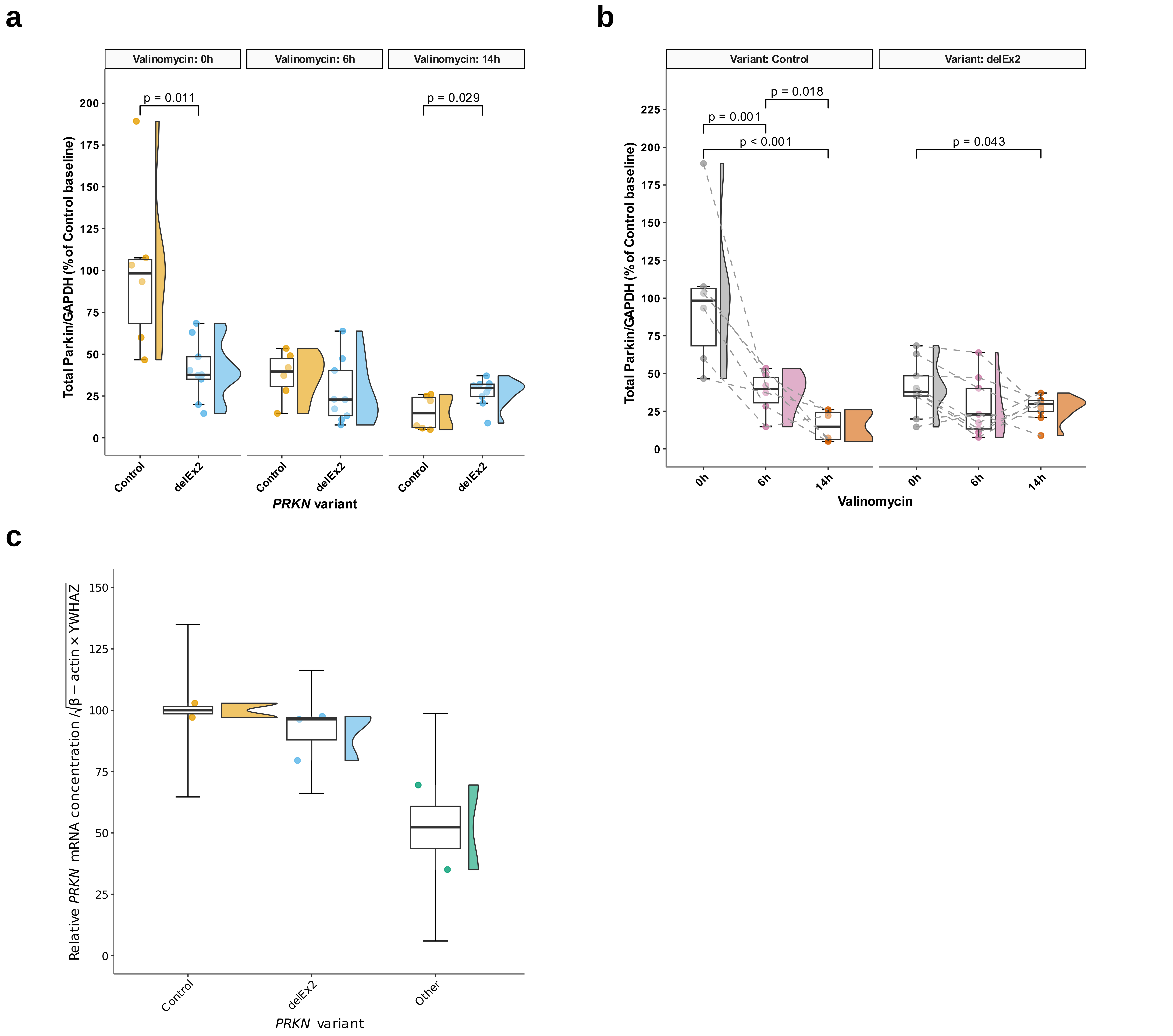


**Supplementary Figure 6:** Total Parkin (full-length Parkin + Parkin$^{\Delta 1-79}$) expression differences between groups (**a**) and expression changes with increased mitochondrial depolarization periods (**b**) in hiPSC-derived midbrain dopaminergic neurons complementary to data in main figure 5. Sample sizes represent a combination of individual patient-derived neurons and independently repeated experiments across three differentiations. The significance threshold was set to p = 0.05. Whiskers extend to the largest and smallest values no further than 1.5 * IQR from the hinge. Post-hoc multiple comparisons via Durbin-Conover tests (**b**) were Holm-adjusted. (**c**) Relative Parkin mRNA expressed in iDNs. Whiskers show error propagated 1.5 * IQR.

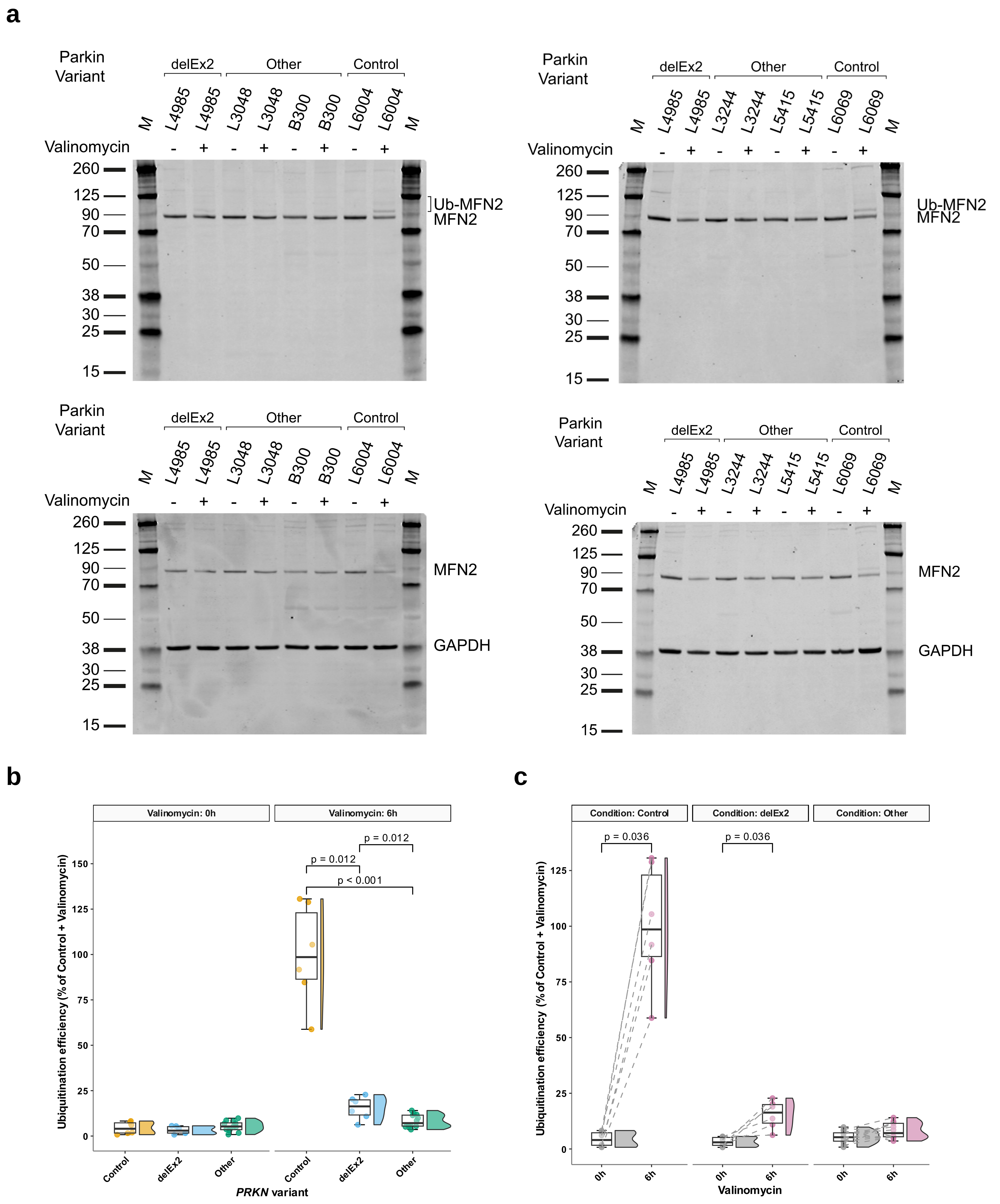


**Supplementary Figure 7: Remaining endogenous MFN2 ubiquitination in biallelic *PRKN*$^{\text{delEx2}}$ patient fibroblasts.**
(**a**) Western blot analysis of MFN2 in 1 µM valinomycin-treated fibroblasts of healthy controls (n = 6), a biallelic *PRKN*$^{\text{delEx2}}$ carrier (n = 6), and carriers of other *PRKN* variants downstream of the internal translation initiation site (n = 12). (**b-c**) Ubiquitination efficiency differences following mitochondrial depolarization between *PRKN* variant carriers and healthy controls (**b**) and change in ubiquitination efficiency between basal and depolarized states (**c**). Sample sizes represent a combination of individual fibroblast lines and independently repeated experiments across three to six passages. The significance threshold was set to p = 0.05. Whiskers extend to the largest and smallest values no further than 1.5 * IQR from the hinge. Post-hoc multiple comparisons via Games-Howell tests (**b**) were Holm adjusted.
